# BRIDGE: a barrier-informed Bayesian Risk prediction model for risk IDentification, trajectory Grouping, and profiling of non-adherencE to cardioprotective medicines in primary care

**DOI:** 10.64898/2026.04.21.26351387

**Authors:** Harvey Jia Wei Koh, Caroline Trin, Zanfina Ademi, Ella Zomer, Danielle Berkovic, Pilar Cataldo Miranda, Brayden Gibson, J. Simon Bell, Jenni Ilomäki, Danny Liew, Christopher Reid, Sean Lybrand, Danijela Gasevic, Arul Earnest, Dragan Gasevic, Stella Talic

**Affiliations:** PharmacoEpidemiology Research Group (PERG), School of Public Health and Preventive Medicine, Monash University, Melbourne, Australia; School of Public Health and Preventive Medicine, Monash University, Melbourne Australia; Health Economics and Policy Evaluation Research (HEPER) Group, Faculty of Pharmacy and Pharmaceutical Sciences, Monash University, Melbourne, Australia; Faculty of Health, Medicine and Behavioural Sciences, University of Queensland, Brisbane, Australia; School of Population Health at Curtin University, Perth, Australia; Centre of Cardiovascular Research and Education in Therapeutics, Monash University; Centre for Global Health, Usher Institute, The University of Edinburgh, Scotland, UK; Center for Learning Analytics, Faculty of Information Technology, Monash University; Faculty of Education and School of Computing & Data Science, The University of Hong Kong, China

## Abstract

**Background:** Non**-**adherence to lipid-lowering therapy (LLT) affects up to half of patients and contributes substantially to preventable cardiovascular morbidity and mortality. Existing measures, such as the proportion of days covered, provide cross-sectional summaries but fail to capture the dynamic patterns of adherence over time. Although group-based trajectory modelling identifies distinct longitudinal adherence patterns, no approach currently predicts trajectory membership prospectively while incorporating patient-reported barriers. We developed BRIDGE, a barrier-informed Bayesian model to predict adherence trajectories and identify their underlying drivers.

**Methods:** BRIDGE incorporates patient-reported barriers as structured prior information within a Bayesian framework for adherence-trajectory prediction. The model was designed not only to estimate which patients are likely to follow different adherence trajectories, but also to generate clinically interpretable probability estimates that help explain why those trajectories may arise and what modifiable factors may be most relevant for intervention.

**Results:** BRIDGE achieved a macro AUROC of 0.809 (95% CI 0.806 to 0.813), comparable to random forest (0.815 (95% CI 0.812 to 0.819)) and XGBoost (0.821 (95% CI 0.818 to 0.824)), two widely used machine-learning benchmarks for structured clinical prediction. Calibration was superior to random forest (Brier score 0.530 vs 0.545; ), and performance was stable across six independent training runs (AUROC SD = 0.003). Incorporating barrier-informed priors improved accuracy by 3.5% and calibration by 5.5% compared to flat priors, showing that incorporation of patient-reported barriers added value beyond electronic medical record data alone. Four clinically distinct adherence trajectories were identified: gradual decline associated with treatment deprioritisation amid polypharmacy (10.4%), early discontinuation linked to asymptomatic risk dismissal (40.5%), rapid decline associated with intolerance (28.8%), and persistent adherence (20.2%). Counterfactual analysis identified trajectory-specific intervention levers.

**Conclusions:** BRIDGE provides accurate and well-calibrated prediction of adherence trajectories while offering clinically actionable insights into their underlying drivers. By integrating patient-reported barriers with routine clinical data, the model supports targeted, mechanism-informed interventions at the point of prescribing to improve adherence to cardioprotective therapies.

**Funding:** MRFF CVD Mission Grant 2017451

*Evidence before this study:* We searched PubMed and Scopus from database inception to December 2025 using the terms “medication adherence”, “trajectory”, “prediction model”, “Bayesian”, “lipid-lowering therapy”, and “barriers”, with no language restrictions. Group-based trajectory modelling has consistently identified three to five adherence patterns across cardiovascular cohorts; however, these applications have been descriptive rather than predictive. Machine-learning models for adherence prediction achieve moderate discrimination but treat adherence as a binary or continuous outcome, thereby overlooking the clinically meaningful heterogeneity captured by trajectory approaches. One prior study applied a Bayesian dynamic linear model to examine adherence-outcome associations, but it did not predict adherence trajectories or incorporate patient-reported barriers. To our knowledge, no published model integrates patient-reported barriers into trajectory prediction.

*Added value of this study:* BRIDGE is, to our knowledge, the first model to incorporate patient-reported adherence barriers as hierarchical domain-informed priors within a Bayesian framework for trajectory prediction. Using 108 predictors derived from routine electronic medical records, the model achieves discrimination comparable to state-of-the-art machine-learning approaches while additionally providing uncertainty quantification, barrier-level interpretability, and counterfactual insights to inform intervention strategies. The identified trajectories differed not only in adherence level but also in switching behaviour, drug-class evolution, and medication burden, suggesting distinct underlying mechanisms of non-adherence that may require tailored clinical responses.

*Implications of all the available evidence:* Each adherence trajectory implies a distinct intervention target: asymptomatic risk communication for early discontinuers (40.5% of patients), proactive tolerability management for rapid decliners, medication simplification for patients with gradual decline associated with polypharmacy, and maintenance support for persistent adherers. By integrating routinely collected clinical data with patient-reported barriers, BRIDGE can be deployed within existing primary care EMR infrastructure to generate actionable, trajectory and patient--specific recommendations at the point of prescribing, helping to bridge the gap between adherence measurement and targeted adherence management.

## INTRODUCTION

Cardiovascular disease (CVD) remains the leading cause of death worldwide, responsible for approximately 18 million deaths annually^1^. In Australia alone throughout 2022 - 2023, approximately 4.5 million people live with CVD, and more than 120 deaths occur daily^2^. Pharmacological therapies, particularly lipid-lowering, antihypertensive, and glucose-lowering agents, are central to cardiovascular risk management; however, their effectiveness depends on sustained medication adherence^3^. Between 15% and 50% of patients fail to take medicines as prescribed, and more than half discontinue lipid-lowering therapy within two years^4^. Non-adherence is estimated to cause 125,000 preventable deaths annually and healthcare costs exceeding €125 billion in Europe and USD 100–300 billion in the United States^5^.

The World Health Organization conceptual framework attributes non-adherence to five interacting domains: patient-related, therapy-related, condition-related, healthcare-system-related, and social and economic-related factors^6^. These include low motivation, poor health literacy, polypharmacy, regimen complexity, adverse effects, and financial stress, among other barriers. Conventional adherence measures, such as the medication possession ratio (MPR) and proportion of days covered (PDC), provide crude, cross-sectional estimates that overlook behavioral change over time^7^.Group-based trajectory modelling (GBTM) has advanced the field by identifying distinct longitudinal adherence patterns^8^; however, existing applications are predominantly descriptive, characterising trajectory groups after the fact rather than predicting membership for a new patient at the point of prescribing. Patient-reported outcome measures (PROMs) capture the contextual and perceptual dimensions of non-adherence, such as intentional versus unintentional behaviour, beliefs about necessity, concerns about side effects, but are rarely integrated into routine clinical workflows^9^.

No existing model combines three elements required for actionable adherence support: (1) longitudinal trajectory prediction capturing heterogeneous ways patients disengage over time, (2) integration of patient-reported barriers to explain *why* patients follow specific trajectories, and (3) transparent, uncertainty-aware predictions with counterfactual reasoning to identify identifiy modifiable intervention targetts. To address these gaps, we developed BRIDGE (**B**arrier-informed Bayesian model for **R**isk **ID**entification, trajectory **G**rouping, and profiling of non-adherenc**E**): a Bayesian hierarchical model for predicting medication adherence trajectories among patients prescribed lipid-lowering therapy using routinely available electronic medical record data and supports nearest-neighbour counterfactual simulation to identify trajectory-specific intervention targets.

## METHODS

### Study design and data sources

We conducted a model development and internal validation study using de-identified, routinely collected general-practice electronic medical record (GP-EMR) data from an Australian primary-care network sourced from IQVIA, spanning January 2013 to March 2023. Data elements included patient demographics, coded diagnoses, prescription records classified by Anatomical Therapeutic Chemical (ATC) code, and laboratory measurements (systolic blood pressure [SBP], LDL cholesterol [LDL-C], and estimated glomerular filtration rate [eGFR]).

We additionally identified patient-reported barriers for interviews and then used a cross-sectional survey to quantify the relative importance of patient-reported barriers by each medication trajectory pattern. Survey data informed the model’s barrier-structured regularisation but were not used as predictors.

Patients were included if they had evidence of incident LLT initiation (ATC prefix C10) during the study period and valid linkage across prescribing, demographic, and laboratory datasets. Patients were excluded if they lacked identifiable LLT initiation or had insufficient data linkage.

A patient was considered adherent if they received a new LLT prescription within 6 months of the previous prescription issue date (sensitivity analysis was conducted within 3 and 9 months to test robustness and consistent with previous studies^10^). In Australian primary care, LLTs are typically prescribed as one original prescription and 5 repeats, meaning 6 dispensings total. If dispensed monthly, this corresponds to roughly 6 months of medication supply. Therefore, EMR-based studies commonly use 6-month prescription renewal as a proxy for adherence or persistence^11,12^.

Ethics approval was obtained under Monash University Human Research Ethics Committee (MUHREC) project number 40627.

### Outcome Definition

The outcome was a four-class adherence trajectory derived from longitudinal prescription dispensing patterns over 36 months or more following first LLT initiation, previously assigned per patient using GBTM^10^ on prescription records:

**G1 Early discontinuation:** Cessation of therapy within the first 6–12 months.

**G2 Rapid decline:** Abrupt cessation after initial adherence period of first 12 months.

**G3 Gradual decline:** Progressive reduction in prescription fills over 12–36 months.

**G4 Persistent adherence (reference group):** Sustained therapy with consistent prescription fill.

Trajectory labels remained static across all prediction time points, representing the long-term adherence phenotype for each patient. Because the model receives updated predictor values at each six-month window (including refill gaps, switching, and biomarker changes that partially reflect the evolving trajectory), BRIDGE functions as a dynamic landmark prediction model: at each observation window, it combines baseline patient characteristics with accumulated treatment-history information to estimate the probability of each long-term trajectory class. Performance metrics therefore reflect this landmark prediction task rather than baseline-only prognostication.

An important methodological consideration is that several time-varying predictors (prescription gaps, switching events, continuity index) may partially reflect the trajectory outcome. To address this, we conducted a baseline-only sensitivity analysis using index-window (0 – 6 months) observations.

### Predictor identification and barrier mapping to medication trajectories

Predictors were identified through a structured, theory-informed process combining: (1) a systematic review of adherence determinants^13^, (2) prior real-world evidence on adherence trajectories^14^, and (3) a cross-sectional survey of Australian patients prescribed cardiovascular medicines^15^.

Patient-reported barriers were identified through: (1) qualitative evidence synthesis on barriers and enablers to medication adherence^13^, (2) a qualitative descriptive study in Australian primary CVD settings^15^, and (3) a consumer-reference-group mapping survey with patients using these medicines. Survey respondents were presented with barrier scenarios mapped to each trajectory pattern and asked to rate which barriers most plausibly drove each pattern. These responses were used to calculate the proportion of respondents endorsing each WHO domain barrier for each trajectory (Supplementary Table S2).

Identified risk factors and barriers were constrained to constructs operationalisable using routinely collected EMR data. Where direct measures were unavailable, empirically reasonable proxy indicators were pre-specified (e.g., geographic remoteness for access barriers, monitoring intensity for engagement, refill gaps for persistence behaviour). Proxy definition were iteratively refined with input from consumer group members to ensure conceptual validity. The full mapping of predictors to WHO domains is provided in Supplementary Table S3.

### Predictor construction

Records were anchored to a patient-level index date defined as the first observed LLT prescription. Predictors were aggregated into non-overlapping six-month windows (windows 1–7; approximately 0–48 months post-initiation). Each patient contributed multiple window-level observations, while the outcome label remained static.

A total of 108 predictors spanned five clinical domains: medication factors (13; LLT subclass exposure, prescription gap days, continuity index, therapy switching, regimen complexity, polypharmacy), healthcare utilisation (9; visit frequency, remoteness classification, monitoring recency, biomarker observation counts), biomarkers (15; SBP, LDL-C, and eGFR estimates with squared terms, imputation indicators, and derived clinical thresholds), demographics (26; age, sex, observation window, smoking status, 17 comorbidity flags, and composite risk scores including comorbidity burden, cardiovascular risk, and metabolic burden), and 45 pre-specified interaction terms capturing clinically plausible effect modification across six categories: demographic interactions (age^2, age x remoteness, sex x age, sex x biomarkers, sex x comorbidity), access and utilisation interactions (remoteness x visit frequency, remoteness x prescription gap, remoteness x biomarker risk), treatment-pattern interactions (switching x gap, switching x continuity, C10 count x polypharmacy, months x switching), comorbidity and risk-score interactions (diabetes x LDL, hypertension x SBP, CKD x eGFR, age x comorbidity, comorbidity x gap, comorbidity x continuity), biomarker interactions (SBP x eGFR, SBP x LDL, eGFR x LDL), and imputation interactions (biomarker value x imputation flag, test recency x imputation flag). Derived binary risk flags (high comorbidity, uncontrolled hypertension, severe CKD, high SBP, high LDL, low eGFR) were included within their respective clinical domains (Supplementary Table S3). Predictor specification was theory-driven, with only pre-defined collinearity pruning applied for model stability.

### Model Specification

BRIDGE is a Bayesian hierarchical multiclass softmax regression with barrier-informed partial pooling^16^. The model predicts the probability of membership in each of the four trajectory classes given a patient predictor vector at any observation window.

Each of the 108 predictors was assigned to one of six WHO-aligned domain groups: personal, medication, condition, health system, social/demographic, and other (Figure 1). The patient-survey-derived barrier proportions were transformed into domain-specific hyperprior scales using 𝜏*_b,c_* ∼ HalfCauchy(0.5 + 2.0 × proportion_!,#_ ) , ensuring that barrier domains more strongly endorsed by patients for a given trajectory received wider prior variances, permitting larger coefficients when supported by data. The linear transformation was chosen so that the baseline scale (0.5) provides moderate regularisation even for domains with zero endorsement, while the slope (2.0) scales the prior width proportionally to endorsement strength; the resulting scale range (0.5-2.5) spans weakly to moderately informative priors. A sensitivity analysis confirmed that alternative scaling functions (slope 1.0 and 4.0) and flat priors (HalfCauchy(0, 2.5) for all domains) produced qualitatively similar coefficient rankings and AUROC (range 0.805–0.811), with the barrier-informed specification yielding the best calibration (Brier score). This hierarchical structure provides partial pooling within barrier domains: groups with many predictors are shrunk appropriately, while barrier domains are allowed to differ by trajectory class.

**Figure 1:**
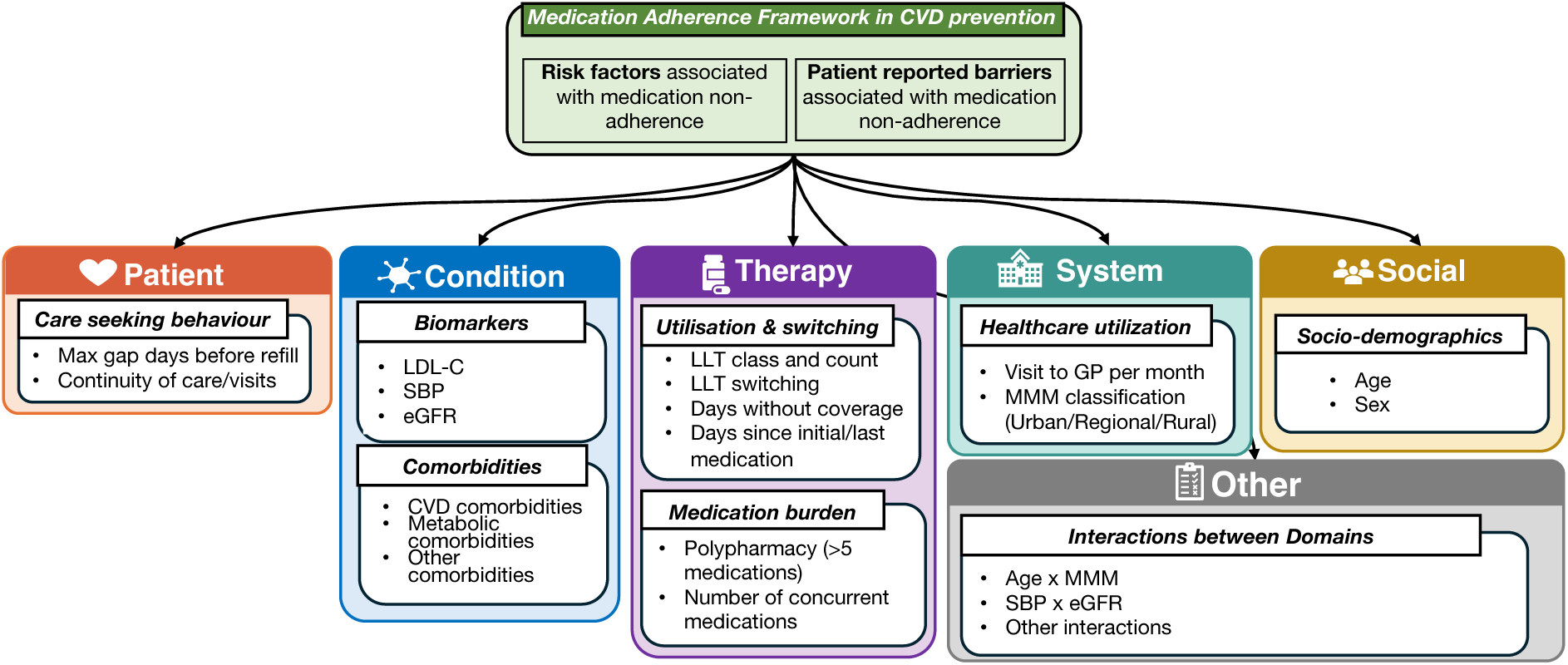
Attributed grouping of each of the major variables to WHO-aligned medication adherence framework groups.

The model uses a categorical likelihood with class-balanced inverse-frequency weighting to address trajectory-class imbalance. Parameters were estimated using stochastic variational inference (SVI) with a diagonal normal variational family (100 000 steps, 500 posterior draws), providing scalable approximate Bayesian inference while preserving uncertainty quantification through the variational posterior (Figure 2). Bayesian model adequacy was assessed through posterior predictive checks and inspection of the learned barrier-domain variance parameters (Supplementary Figure S4). Full mathematical specification of the likelihood, priors, and hyperpriors is provided in the Supplementary Methods 3.

**Figure 2.**
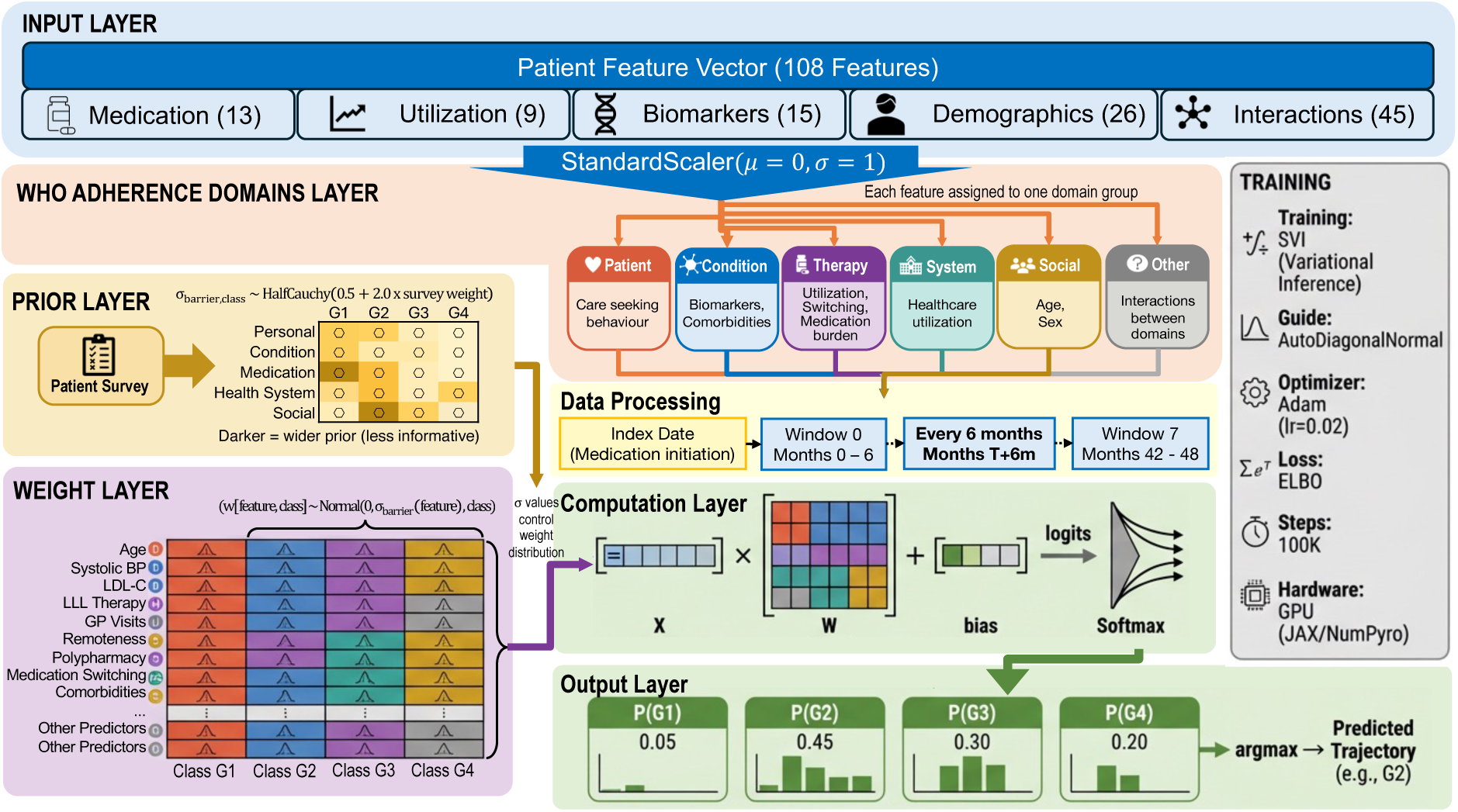
BRIDGE model architecture: A 108-dimensional patient feature vector spanning five clinical domains: Medication (13), Utilisation (9), Biomarkers (15), Demographics (26), and Interactions (45). The feature vector is standardised (mean = 0, SD = 1) and passed through a prior layer that assigns clinician-informed regularisation weights according to WHO adherence barrier categories (Patient, Condition, Therapy, System, Healthcare Access, Social). Observations are constructed at six-month windows from treatment initiation (Window 0 at index date through Window 7 at 42-48 months). The computation layer applies Bayesian multinomial logistic regression, producing class probabilities via softmax across four trajectory groups: G1 (Gradual Decline), G2 (Early Discontinuation), G3 (Rapid Decline), and G4 (Persistent Adherence). The trajectory with maximum predicted probability is assigned as the final classification.

### Model fitting and validation

The dataset was partitioned into patient-level train (𝑛 = 43 245) and test (𝑛 = 7 636) splits stratified by trajectory group (85/15). To ensure robustness of results, the full pipeline was repeated six times with different random seeds, primary results are reported as mean ± SD across seeds, with bootstrap 95% confidence intervals from the representative seed (seed 42). Metrics included area under the receiver operating characteristic curve (AUROC), accuracy, and Brier score (measuring probability calibration). Bootstrap CIs were computed at the observation-window level on the held-out test set ( = 28 107 windows from 7 636 patients).

BRIDGE was compared against two machine-learning baselines, random forest and XGBoost, two state-of-the-art machine learning algorithms for structured data^17^, trained on the same train/test split across all six seeds. A sensitivity analysis using five-fold cross-validation compared BRIDGE with barrier-informed priors against a model with flat (uninformative) priors to quantify the contribution of the barrier hierarchy.

### Trajectory group characterisation

To move beyond aggregate performance metrics and understand *what* the model reveals about patients, we conducted a post-hoc characterisation of the four trajectory groups in the dataset. Per-patient features were aggregated across observation windows (e.g., maximum follow-up duration, total switching events, mean visit count). Additional longitudinal features were derived: early switching rate (0–6 month window), retention at 12 months, LDL-C trajectory (last minus baseline LDL-C), and LLT persistence (ratio of last to baseline number of C10 drug categories).

Switching-pattern analysis classified each switch event as within-class (same C10 subclass composition, e.g., statin-to-statin dose change), between-class (addition or substitution of a different drug class), or baseline (occurring in the first six-month window). Longitudinal drug-class composition was tracked across all eight-time windows.

Group differences were assessed using Kruskal–Wallis tests (continuous variables) and 𝜒^$^ tests (binary variables), with Bonferroni correction for multiple comparisons. Effect sizes were quantified using Cohen’s 𝑑 (continuous) and Cramér’s 𝑉 (binary).

### Counterfactual intervention analysis

To identify modifiable intervention levers for each non-adherent trajectory, we conducted a three-part counterfactual analysis on the held-out test set using all available observation-window timepoints, restricted to correctly classified observations (i.e., patients whose predicted trajectory matched their actual group assignment, ensuring that counterfactual shifts are interpreted from a valid baseline).

First, we decomposed BRIDGE’s predictions into WHO barrier-domain contributions by computing the mean absolute feature×weight product for each predictor, aggregated by domain. Interaction terms (44 features in the “other” domain) were assigned post-hoc to their constituent WHO domains using pre-specified fractional weights (e.g., age x MMM was split 0.5 personal, 0.5 medication), ensuring that all predictive signal was attributed to clinically interpretable barrier domains. The difference in domain contributions between each non-adherent group and the persistent-adherence (G4) reference was computed to identify domains where each group carried excess or deficit barrier load.

Second, we estimated population-level probability shifts using nearest-neighbour (NN) counterfactual matching. For each non-G4 patient, we identified the 𝑘 = 10 nearest G4 (persistent adherence) neighbours using Euclidean distance on 21 standardised non-modifiable features (age, sex, remoteness classification, 17 comorbidity indicators, and months since treatment initiation). Nearest-neighbour is chosen to specifically identify closest possible patient profiles from the persistent group to compare to non-adherence groups. For each intervention scenario, the patient’s modifiable base features were replaced with the mean values of their G4 neighbours, and interaction terms were recomputed from the resulting feature vector to maintain internal consistency; temporal interaction terms involving months since initiation were frozen at their original values to avoid post-treatment bias. Six intervention scenarios were tested: (i) prescription timeliness (maximum gap days, days since last dispensing), (ii) medication switching (therapy switching, number of C10 categories), (iii) regimen simplification (polypharmacy flag, unique ATC codes), (iv) blood pressure control (SBP and derived features), (v) LDL cholesterol control (LDL and derived features), and (vi) smoking cessation. A greedy “all interventions combined” scenario was constructed by adding levers sequentially in order of decreasing individual Δ𝑃(G4) , retaining only those that improved the cumulative effect; this identifies the optimal intervention bundle for each trajectory while accounting for non-linear interaction effects that can cause combined interventions to be less effective than the sum of their parts.

Third, we conducted a medication simplification analysis targeting six regimen-change scenarios (reducing total medicine types, removing polypharmacy, consolidating to single LLT class, lipid-lowering polypill, statin-only therapy, and deprescribing two medicines). All scenarios respected a per-patient comorbidity-aware minimum (one medicine for LLT plus one per major comorbidity) and were evaluated at both the population level and within the polypharmacy subgroup (≥5 concurrent medications). Full scenario definitions, comorbidity floor rules, and medicine-count operations are detailed in the Supplementary Methods 3.

### Partial dependence analysis

To visualise how individual clinical factors influence the predicted probability of persistent adherence over time, we constructed partial-dependence grids. For each factor of interest, all other predictors were held at their test-set median values while the target factor and months since initiation were varied across their clinically relevant ranges. Interaction terms involving the swept variables (e.g., comorbidity × gap, switching × continuity, C10 categories × months) were recomputed at each grid point to maintain internal consistency. Five factors were selected for their clinical relevance and modifiability: comorbidity count, number of concurrent C10 categories, therapy switching, total unique medications, and GP visit frequency.

## RESULTS

### Cohort characteristics

The study included 50 881 adults initiating LLT across 43 245 training and 7 636 test patients (Figure 3). Mean age was 62.9 years (SD 13.5), 53% were male, and 74% resided in major cities (Modified Monash Model category 1). Statins were the predominant therapy (90%), with uncommon use of fibrates, bile-acid sequestrants, and other agents (<5% each). Polypharmacy (≥5 concurrent medications) affected approximately 10% of patients. Four distinct trajectories were observed with consistent distribution across cohorts: gradual decline (10%), early discontinuation (41%), rapid decline (29%), and persistent adherence (20%; Table 1).

**Figure 3:**
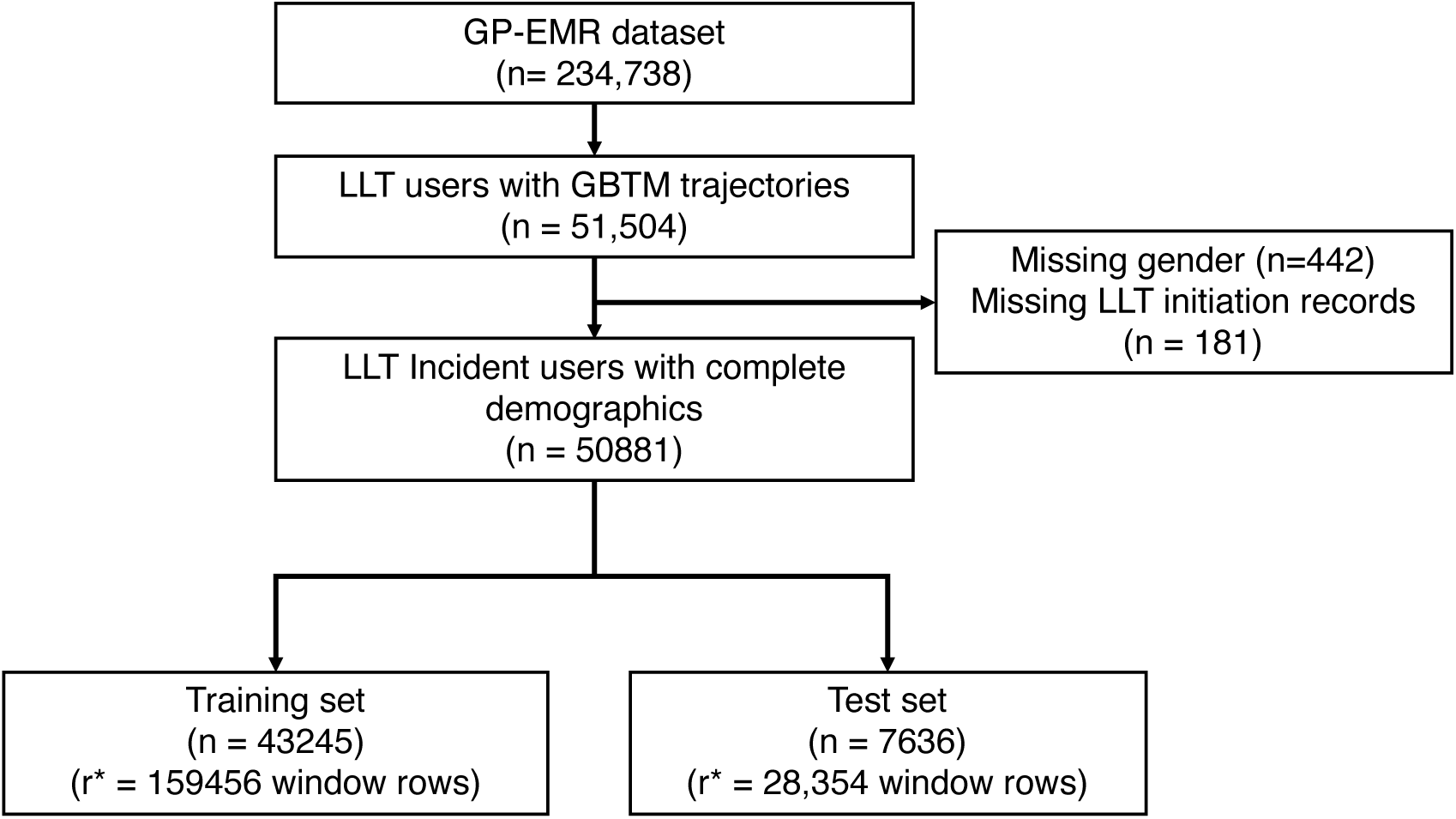
Data flow, inclusion and exclusion criteria and final derived training and test sets.

**Table 1:** Patient characteristics of train/test datasets.

| Variable | Train (n = 43 245) | Test (n = 7 636) |
| --- | --- | --- |
| Age, years, mean (SD) | 62.9 (13.5) | 62.8 (13.6) |
| **Remoteness (MMM), n (%)** |  |  |
| Urban | 31 794 (73.5%) | 5 602 (73.4%) |
| Regional | 6 436 (14.9%) | 1 118 (14.6%) |
| Rural | 5 015 (11.6%) | 916 (12.0%) |
| **Trajectory group, n (%)** |  |  |
| G1 – Gradual decline | 4 508 (10.4%) | 797 (10.4%) |
| G2 – Early discontinuation | 17 539 (40.6%) | 3 096 (40.5%) |
| G3 – Rapid decline | 12 464 (28.8%) | 2 201 (28.8%) |
| G4 – Persistent adherence | 8 734 (20.2%) | 1 542 (20.2%) |
| **Medications, n (%)** |  |  |
| Statin use | 38 925 (90.0%) | 6 871 (90.0%) |
| Fibrate use | 1 888 (4.4%) | 353 (4.6%) |
| Bile acid sequestrant use | 205 (0.5%) | 44 (0.6%) |
| Other lipid-lowering use | 1 288 (3.0%) | 232 (3.0%) |
| Statin + other combo use | 1 849 (4.3%) | 354 (4.6%) |
| Other lipid-lowering combo use | 890 (2.1%) | 165 (2.2%) |
| Lipid-lowering drug count | 1.042 (0.210) | 1.050 (0.233) |
| Number of LLT agents used | 1.042 (0.210) | 1.050 (0.233) |
| Polypharmacy (≥5 meds) | 3 981 (9.2%) | 698 (9.1%) |
| Switched therapy (within window) | 1 385 (3.2%) | 302 (4.0%) |
| Average medication count | 2.300 (1.508) | 2.303 (1.494) |
| **Comorbidities, n (%)** |  |  |
| Alcohol-related liver disease | 12 (0.01%) | 2 (0.01%) |
| Coronary heart disease / angina | 2 625 (6.1%) | 480 (6.3%) |
| Atrial fibrillation | 1 223 (2.8%) | 244 (3.2%) |
| Coronary artery bypass graft | 845 (2.0%) | 143 (1.9%) |
| Chronic kidney disease | 1 098 (2.5%) | 180 (2.4%) |
| Peripheral artery disease | 365 (0.8%) | 69 (0.9%) |
| Diabetes mellitus | 6 871 (15.9%) | 1 159 (15.2%) |
| Familial hyperlipidaemia | 89 (0.2%) | 14 (0.2%) |
| Heart failure | 871 (2.0%) | 157 (2.1%) |
| Hyperlipidaemia | 11 000 (25.4%) | 1 951 (25.6%) |
| Hypertension | 10 852 (25.1%) | 1 910 (25.0%) |
| Myocardial infarction | 746 (1.7%) | 140 (1.8%) |
| Non-alcoholic liver disease | 35 (0.1%) | 13 (0.2%) |
| Obesity | 134 (0.3%) | 24 (0.3%) |
| Percutaneous coronary intervention | 7 (0.0%) | 2 (0.0%) |
| Smoking status | 5 171 (12.0%) | 897 (11.7%) |
| Stroke / TIA | 721 (1.7%) | 135 (1.8%) |
| Viral hepatitis | 13 (0.01%) | 1 (0.01%) |
| **Biomarkers and composites** |  |  |
| eGFR, mL/min/1.73m<sup>2</sup>, mean (SD) | 75.269 (12.146) | 75.370 (11.980) |
| LDL cholesterol, mmol/L, mean (SD) | 2.537 (0.619) | 2.534 (0.631) |
| SBP, mmHg, mean (SD) | 131.124 (8.826) | 131.100 (8.700) |
| Comorbidity count, median (IQR) | 1 (0-2) | 1 (0-2) |
| High comorbidity ($\geq 3$), n (%) | 4 274 (9.9%) | 762 (10.0%) |

### Model performance

BRIDGE achieved macro-averaged AUROC 0 . 809 (95% CI 0 . 806–0 . 813), statistically equivalent to random forest (0.815, 0.812–0.819) and comparable to XGBoost (0.821, 0.818–0.824; Table 2). BRIDGE achieved superior probability calibration over random forest (Brier score 0⋅530, 0⋅526–0⋅535 vs 0⋅545, 0⋅541–0⋅549; non-overlapping CIs) and comparable calibration to XGBoost (0⋅517, 0⋅512–0⋅522).

**Table 2:** Model performance on the held-out test set.

| Model | AUROC (95% CI) | Accuracy (95% CI) | Brier score (95% CI) |
| --- | --- | --- | --- |
| BRIDGE (Bayesian) | 0.808 (0.805–0.811) | 0.585 (0.580–0.590) | 0.531 (0.528–0.534) |
| Random forest | 0.815 (0.812–0.818) | 0.583 (0.578–0.588) | 0.548 (0.544–0.551) |
| XGBoost | 0.820 (0.817–0.823) | 0.597 (0.592–0.602) | 0.518 (0.514–0.521) |

Bootstrap 95% confidence intervals from 5000 resamples. AUROC = area under the receiver operating characteristic curve. Lower Brier scores indicate better probability calibration. Results were stable across six independent training runs (AUROC SD < 0.01 for all models).

For reference, the expected Brier score under random prediction for a four-class problem is 0⋅75 (1 − 1/𝐾). Balanced accuracy (mean per-class recall) was 0⋅53 for BRIDGE, reflecting strong discrimination of G2 (recall 0⋅71) and G4 (0⋅80) but poor recall for the smallest class G1 (0⋅09, 𝑛 = 797 in the test set). Overall accuracy (0⋅59) modestly exceeds the majority-class baseline (0⋅41 if always predicting G2), but the model value lies in probability calibration and trajectory-specific risk stratification rather than hard classification. Results were stable across five independent training runs (AUROC SD < 0⋅01 for all models; Table 2). Calibration and subgroup fairness are in Supplementary Figures S1–S8.

Per-class discrimination varied by trajectory (Figure 4): G2 (early discontinuation) was best discriminated (AUROC 0⋅941), followed by G4 (persistent adherence, 0⋅827), G3 (rapid decline, 0⋅763), and G1 (gradual decline, 0⋅707). Subgroup fairness analysis showed consistent performance across age groups, sex, and geographic remoteness, with no clinically meaningful degradation in any subgroup (Supplementary Figures S2–S3). Posterior predictive checks confirmed that the model’s predicted class distributions closely recovered the observed prevalences for all four trajectories (Supplementary Figure S4), and the learned barrier-domain variance parameters were consistent with the hypothesised barrier profiles, with therapy-related barriers exhibiting the largest posterior variance for G2 and socioeconomic factors for G3 and G4.

**Figure 4.**
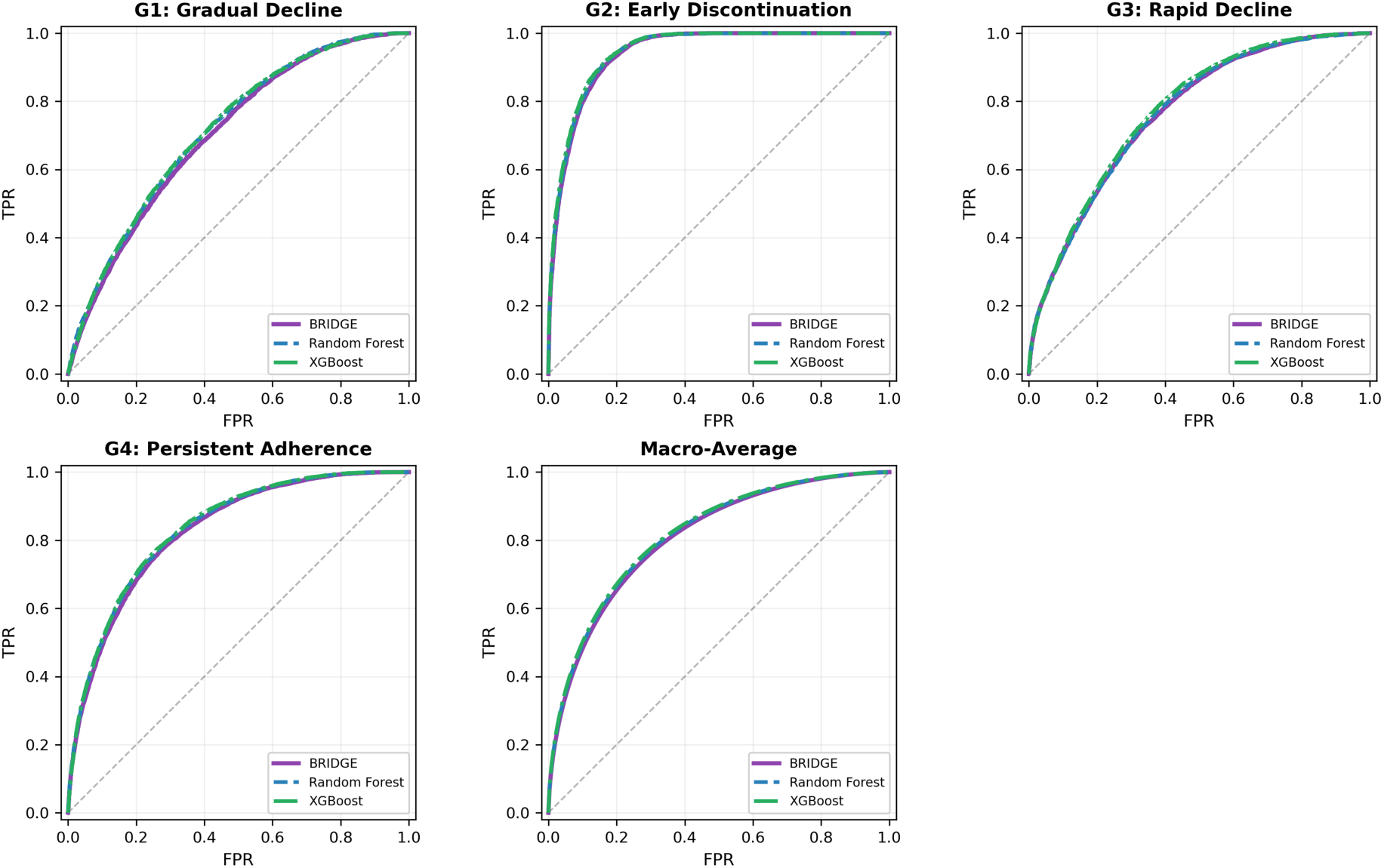
Held-out test-set model comparison: BRIDGE, random forest, and XGBoost were all trained on the full training cohort and evaluated on the fixed test cohort. Macro AUROC (95% CI): BRIDGE 0.81 (0.81–0.81), random forest 0.82 (0.81–0.82), and XGBoost 0.82 (0.82–0.82). Results were stable across six random seeds (SD < 0.01 for all models).

### Prediction Stability

Retraining five times with different seeds confirmed stability: macro AUROC ranged from 0.80 to 0.81 (mean 0.81, SD < 0.01), with per-class AUROCs similarly stable (G4 mean 0.83, SD = 0 . 004; Supplementary Table S1). The barrier-informed model showed consistent improvements over flat priors in accuracy (59% vs 55%) and calibration (Brier 0.53 vs 0.56).

### Trajectory group characterisation

Three non-adherence trajectories were identified (G1-G3), each exhibiting distinct demographic, clinical, and treatment profiles when compared with the persistent-adherence reference group G4 (Figure 5).

**Figure 5:**
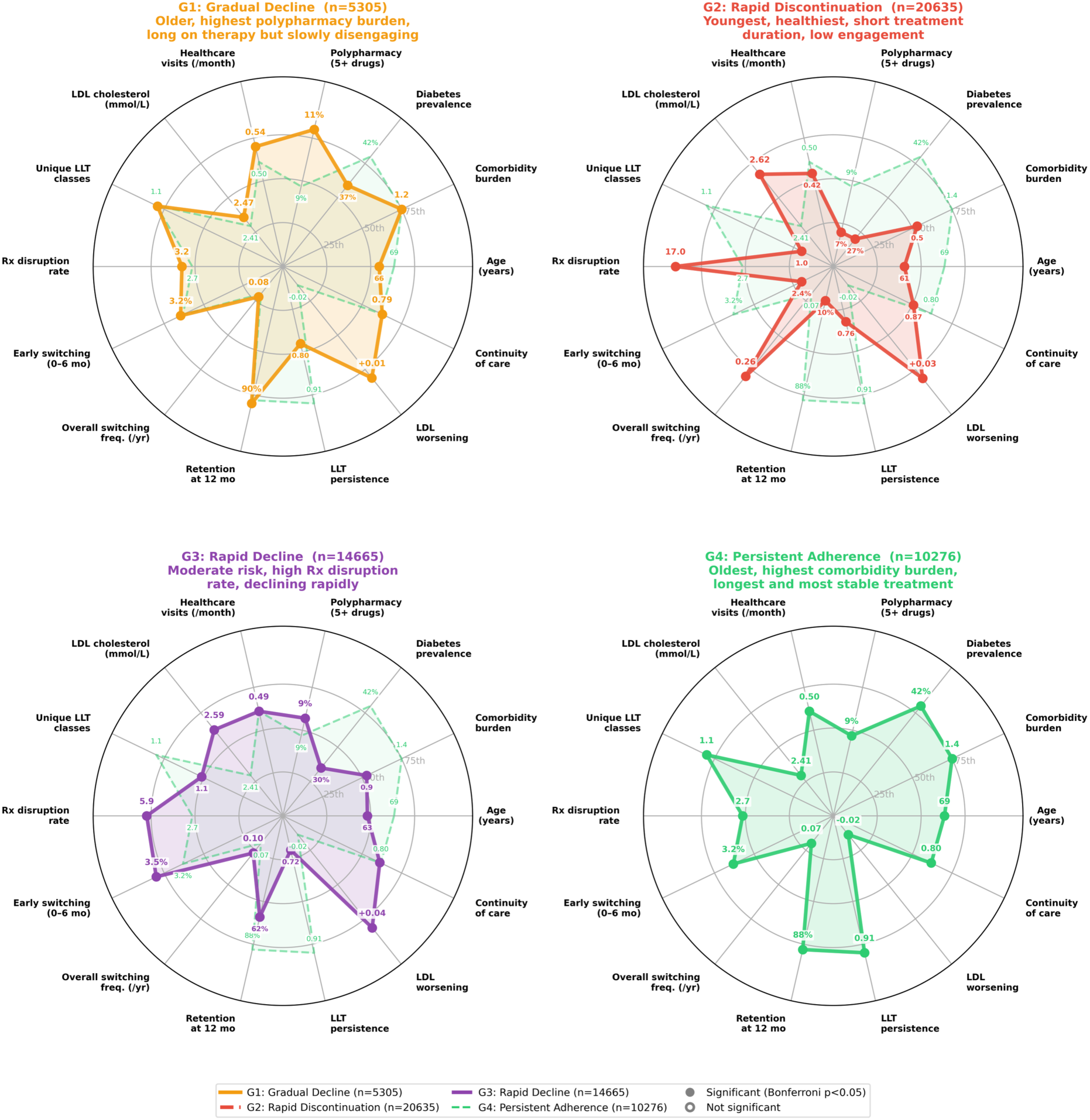
Trajectory group archetype profiles compared with the persistent-adherence (G4) reference. Radar plots showing per-patient aggregated features, normalised to percentile rank within the full cohort distribution. Each panel shows one non-adherent group (solid coloured line) against G4 (dashed green line), with G4 shown standalone in the bottom-right panel. Solid circular markers indicate features with significant omnibus group differences (Bonferroni-corrected 𝑝 < 0.05, Kruskal–Wallis or 𝜒^2^); hollow markers indicate non-significant differences. Annotated values show raw group means

**G1: Gradual decline (10.4%, n=5,305).** Gradual decliners were clinically similar to persistent adherers (mean age 66 vs 69 years, comparable hypertension prevalence) but had the highest polypharmacy of any group (10.6% vs 8.6% in G4) despite fewer comorbidities, suggesting greater medication complexity. Statin use eroded gradually from 91% to 54% over four years, yet healthcare engagement remained high (0.55 visits/month, 89% retained at 12 months). Among the 11% who switched therapy, switching was predominantly between-class escalation rather than dissatisfaction. This combination of sustained engagement, high medication burden, and slow disengagement over 33 months is consistent with a treatment-fatigue hypothesis in which LLT may be deprioritised amid competing medication demands.

**G2: Early discontinuation (40.5%, n=20,635).** The largest group was also the youngest (mean age 61 years) and healthiest, with the lowest comorbidity burden (0.5 ± 0.9), lowest polypharmacy (7.0%), and poorest lipid control (LDL-C 2.6 mmol/L). Only 11% were retained at 12 months; prescription disruption was extreme (16 days/month of follow-up). These patients simply stopped without trialling alternatives: only 2.7% ever switched, and 99% of those events occurred at baseline. The absence of switching suggests intolerance was unlikely the primary driver. This pattern of early, complete disengagement is consistent with low perceived treatment necessity in the context of asymptomatic cardiovascular risk.

**G3: Rapid decline (28.8%, n=14,665).** Rapid decliners were middle-aged (mean 63 years) with intermediate comorbidity burden (0.9 ± 1.1) and the lowest LLT persistence of any group (0.71). The defining feature was early, unsuccessful switching: among the 7.0% who switched, the median first switch was at 6 months (the earliest of any group), with 47% at baseline, yet statin use still eroded from 89% to 24% by 36 months. This pattern of trialling alternatives followed by progressive discontinuation is consistent with an intolerance-driven dropout.

**G4: Persistent adherence (20.2%, n=10,276).** Persistent adherers were the oldest (mean age 69 years) with the highest clinical burden (diabetes 42%, CKD 8.8%), lowest LDL-C (2.4 mmol/L, improving over time), and the lowest prescription disruption (2 . 8 days/month). Polypharmacy was moderate (8 . 6%), notably lower than G1 despite higher comorbidity, suggesting better-rationalised prescribing. This was the only group in which treatment complexity increased over time: ezetimibe/PCSK9 inhibitor use rose from 3.8% to 5.3%, with switching dominated by within-class optimisation (44%) and escalation to combination therapy (48%), consistent with guideline-concordant step-up care.

### Counterfactual intervention analysis

We performed a three-part counterfactual analysis across all observation-window timepoints, restricted to correctly classified observations ( 𝑛 = 50 849: G1 = 2 410, G2 = 22 467, G3 = 25 972; Figure 6). For each non-adherent group, we decomposed barrier contributions (Panel A), estimated probability shifts under nearest-neighbour counterfactual matching (Panel B) and evaluated medication simplification scenarios (Panel C). All medicines-count reductions respected a per-patient comorbidity-aware floor. These are model-based projections, not causal estimates, and the effect sizes are modest (largest combined shift: +4.5 pp for P(G4) in G2).

**Figure 6:**
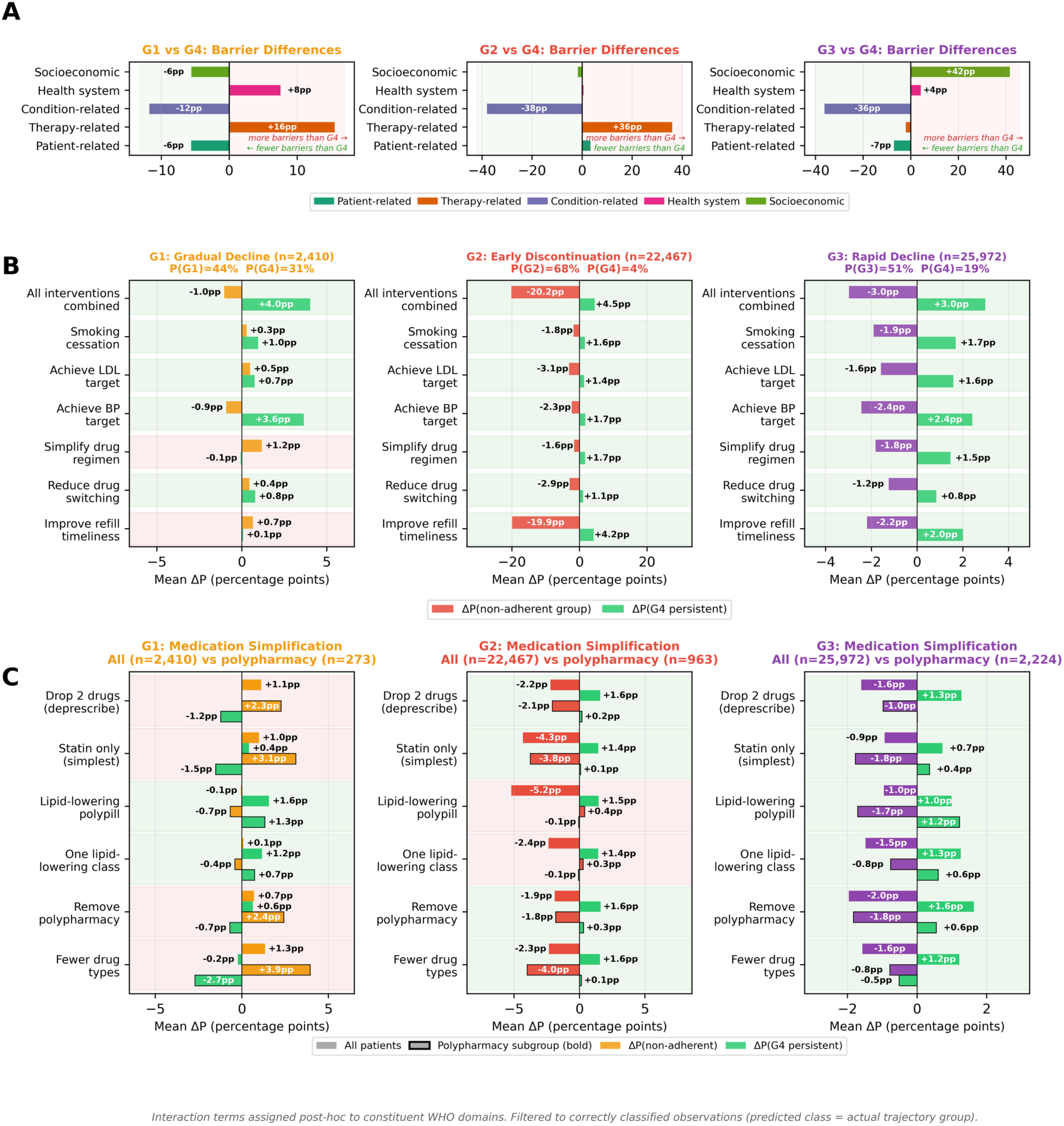
Counterfactual intervention analysis (patient-level, all timepoints, correctly classified observations, 𝑛 = 50 849). Panel. **A**: Barrier-domain contributions relative to the persistent-adherence (G4) reference. **Panel B**: Probability shifts under nearest-neighbour counterfactual matching (𝑘 = 10 G4 neighbours on 21 non-modifiable features; base modifiable features substituted, cross-interactions recomputed). Six intervention scenarios plus a greedy “all combined” scenario. **Panel C**: Medication simplification analysis across six regimen-change scenarios. Faded bars show population-level effects; bold-outlined bars show the polypharmacy subgroup (≥5 concurrent drugs). All drug-count reductions respect a per-patient comorbidity-aware floor. Full scenario definitions are in the Supplementary Methods

**G1 Gradual decline**: G1 predictions were driven by condition-related (51%) and therapy-related (25%) barriers, with excess therapy-related load (+15 pp vs G4). BP control was the dominant lever (+3.6 pp for P(G4)), with smoking cessation (+1.0 pp) and medicine switching (+0.8 pp) contributing to a combined + 4 . 0 pp for P(G4). Among polypharmacy patients ( 𝑛 = 273), reducing total medicine count paradoxically worsened adherence predictions (+2.3 pp for P(G1)), reflecting disease-severity-driven persistence in this multimorbid subgroup. The only beneficial simplification strategies were statin-only therapy (+1.1 pp for P(G4)) and the lipid-lowering polypill (+0.7 pp), which reduce regimen complexity without lowering total medicine count.

**G2 Early discontinuation**: G2 predictions were dominated by therapy-related barriers (+36 pp vs G4), consistent with prescribing-level disengagement. Prescription timeliness was the single most impactful lever (−19.9 pp for P(G2), +4.2 pp for P(G4)), with BP control (+1.7 pp for P(G4)) and LDL-C control (+1.4 pp) as secondary levers. Greedily combining all beneficial interventions reduced P(G2) by 20.2 pp and increased P(G4) by +4.5 pp – the largest combined shift of any group. Medication simplification added further benefit: statin-only therapy reduced P(G2) by 3. 0 pp at the population level, with removing polypharmacy shifting P(G2) by −1.8 pp among polypharmacy patients (𝑛 = 963). These findings suggest that G2 patients, who disengage early without trialling alternatives, are the primary candidates for proactive prescription monitoring and regimen simplification.

**G3 Rapid decline:** G3 was driven by socioeconomic barriers (+42 pp vs G4), with minimal therapy-related contribution (8%). BP control was the strongest individual lever (+2.4 pp for P(G4)), followed by prescription timeliness (+2.0 pp) and smoking cessation (+1.7 pp), with all beneficial interventions combined yielding + 3 . 0 pp. Among polypharmacy patients (𝑛 = 2 224), removing polypharmacy shifted P(G3) by −1.8 pp and P(G4) by +0.6 pp. The modest effect sizes are consistent with socioeconomic barrier dominance: clinical interventions alone have limited reach when non-clinical factors drive disengagement, underscoring the need for complementary social and structural supports.

### Intervention timing: window-stratified counterfactual analysis

To identify when interventions are most effective, we repeated the nearest-neighbour counterfactual analysis separately at each six-month observation window, nearest-neighbour is used to choose a patient with the closest risk profile (Supplementary Table S6; Supplementary Figure S7). Intervention effects grew monotonically with time on therapy for all three non-adherent groups, consistent with progressive behavioural divergence from persistent adherers creating larger scope for modifiable-feature substitution at later windows. When aggregated into early (0–12 months), mid (18–30 months), and late (36–42 months) treatment phases, the sample-size-weighted combined Δ𝑃(G4) increased from +3.1 pp to +4.5 pp to +7.3 pp for G1, from +4.5 pp to +6.6 pp to +10.4 pp for G2, and from +2.8 pp to +5.8 pp to +9.7 pp for G3.

Lever-specific patterns differed across trajectories. For G1 (gradual decline), BP control was the dominant lever from 12 months onward (+2.5 pp at 12 months, rising to +6.4 pp at 42 months), while prescribing-level levers (prescription timeliness, switching) were ineffective or counterproductive during early windows. For G2 (early discontinuation), prescription timeliness was the strongest individual lever across all windows but was especially dominant at early windows (+3.6 pp at baseline, representing 91% of the combined effect), consistent with prescribing disengagement as the primary mechanism. For G3 (rapid decline), all levers showed similar growth trajectories, with prescription timeliness and regimen simplification converging at late windows (+8.4 pp and +7.7 pp respectively at 42 months). These trajectory-specific temporal profiles have implications for intervention design: G2 patients should be targeted early with prescription monitoring, while G1 patients benefit most from sustained biomarker management over time.

### Partial-dependence analysis

To understand how clinical factors interact with time to shape adherence persistence, we constructed partial-dependence grids for seven key dimensions (Figure 7; extended version in Supplementary Figure S5).

**Figure 7:**
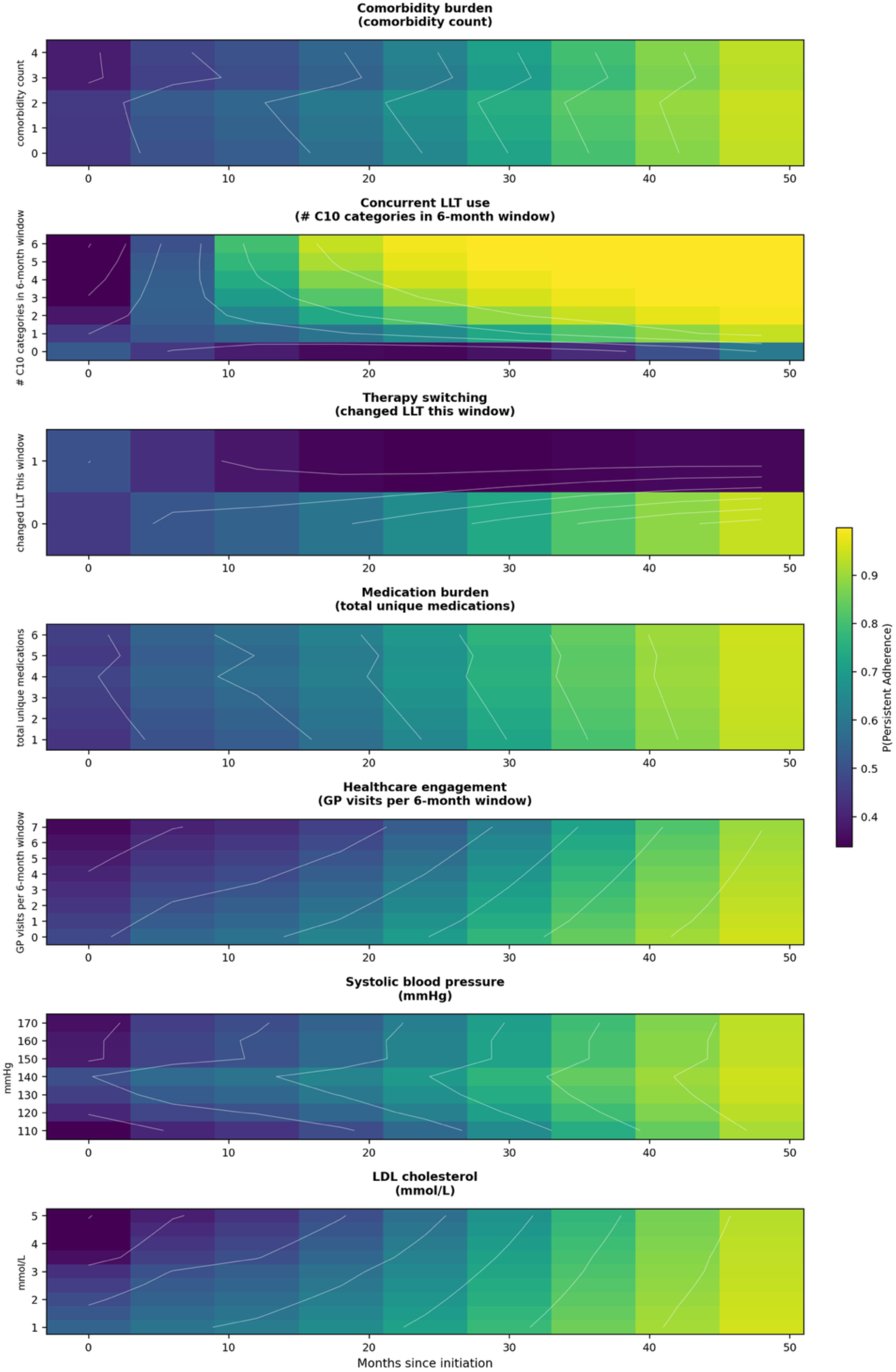
Partial-dependence analysis predicted probability of persistent adherence (G4) as a function of time since initiation and key clinical factors. Each panel shows how 𝑃(Persistent Adherence) varies across months since LLT initiation (x-axis) and levels of one clinical factor (y-axis), holding all other features at their test-set median. Colour represents 𝑃(G4) on a shared viridis scale (2nd–98th percentile); white contour lines highlight iso-probability gradients. **Comorbidity burden**: higher comorbidity count is associated with greater 𝑃(G4), consistent with disease-severity-driven motivation. **Concurrent LLT use**: patients prescribed multiple C10 categories have markedly higher persistence, particularly beyond 12 months. **Therapy switching**: switching therapy within a 6-month window approximately halves the predicted persistence probability, the single strongest modifiable risk signal. **Medication burden**: total unique medication count shows a modest positive gradient reflecting polypharmacy as a proxy for disease complexity and healthcare engagement. **Healthcare engagement**: higher GP visit frequency is associated with greater persistence, particularly in the first 24 months. **Systolic blood pressure**: higher SBP is associated with lower P(G4), with a 20-percentage-point difference between 110 and 170 mmHg at 24 months. **LDL cholesterol**: higher LDL shows a strong negative gradient, falling from approximately 0 at 1 mmol/L to 0 at 5 mmol/L, consistent with the counterfactual finding that biomarker optimisation is a key modifiable lever.

The strongest signals fell into two categories: treatment-pattern factors that are themselves markers of engagement, and biomarker factors that are clinically modifiable. Among treatment-pattern factors, therapy switching was the single strongest risk signal: at 24 months, switching LLT halved 𝑃(G4) from 0.69 to 0.31, consistent with the intolerance-driven dropout pattern observed in G3. Conversely, the number of concurrent C10 categories showed the steepest positive gradient (𝑃(G4) rising from 0.39 with no active LLT to 0.98 with six categories at 24 months), reflecting active therapeutic optimisation. Healthcare engagement (GP visits) showed a moderate gradient (𝑃(G4) from 0.58 with zero visits to 0.78 with seven), while comorbidity burden and total medicine count had modest positive effects consistent with disease-severity-driven persistence.

Among modifiable biomarkers, uncontrolled BP and LDL-C cholesterol were both associated with markedly lower persistence: at 24 months, 𝑃(G4) dropped by 20 pp between 110 and 170 mmHg SBP, and by 30 pp between 1.0 and 5.0 mmol/L LDL. These gradients corroborate the intervention simulation, where biomarker optimisation was the most effective lever for G1 and G3. Uncontrolled biomarkers may reflect both inadequate pharmacotherapy and disengagement from care, suggesting a bidirectional relationship between disease control and adherence persistence. Finally, 𝑃(G4) rose from 0.47 at month 0 to 0.95 at month 48, reflecting both genuine survivorship and the landmark prediction design (see Methods); the clinical implication is that the model’s predictive value is highest at early windows, when adherence support decisions are most actionable.

## DISCUSSION

### Principal findings and interpretation

In a large, real-world cohort of Australian primary-care patients initiating LLT, BRIDGE predicted membership across four established adherence trajectories and characterised both trajectory-specific and shared barrier profiles using routinely available EHR data. To our knowledge, this is the first adherence risk prediction model that combines 1) group-based trajectory modelling and a temporal Bayesian variational inference risk prediction model that is on par with state-of-the-art ML models. BRIDGE has demonstrated very good discrimination and class-wise calibration, performing comparably to Random Forest and similarly to XGBoost, while retaining interpretability through its barrier-informed hierarchical priors. 2) Combines explainability of predictions through qualitative barrier priors via the WHO domain and provide counterfactuals to suggest potential interventions for better adherence. In primary care settings, where uptake depends on trust, calibration, and explanation, this balance may be as important as *minimal* incremental gains in model discrimination performance. These findings align with the growing emphasis on context, credibility, and clinical utility in decision support within digital medicine^18^.

### How this research compares with other studies

Most adherence-prediction studies have relied on static summary metrics, typically PDC/MPR, or have characterised longitudinal patterns retrospectively using GBTM. Although PDC is the preferred claims-based adherence metric in quality programmes, it collapses dynamics over time and can mask heterogeneous disengagement patterns. By contrast, GBTM studies across therapeutic areas (including statins) consistently identify four to six archetypal trajectories, commonly high/persistent adherence, gradual decline, rapid decline, and early discontinuation, supporting the face validity of the trajectory structure used here. Simulation and empirical evidence further suggest that GBTM better captures heterogeneity than PDC for understanding longitudinal behaviour^19–21^.

However, most trajectory studies remain descriptive^8,14^, classifying patients after follow-up rather than prospectively predicting trajectory membership at or near initiation. Where prediction has been attempted, models have typically used standard ML classifiers trained on claims or EHR data, often without incorporating behavioural structure and with inconsistent reporting of calibration and external validation^22^. In addition, much of the existing adherence-prediction literature focuses on patient-level clinical characteristics or dispensing-derived behaviour, while giving relatively little attention to prescriber-level mechanisms that may influence whether treatment is started, continued, or deprioritised. Factors such as the timing of planned review, the urgency attached to biomarker follow-up, and the clinician’s verbal framing of medicine necessity are likely to affect adherence behaviour, yet are rarely represented in routine modelling datasets^23,24^. This omission may be particularly relevant for preventive therapies, where the perceived importance of treatment is strongly shaped by how risk and benefit are communicated. In parallel, there is a literature on predicting binary or next-event adherence using ML and deep learning (e.g., LSTM), but these approaches typically target short-horizon events rather than long-term trajectory phenotypes and often prioritise predictive performance over interpretability^25,26^. BRIDGE combines dynamic adherence patterns for prediction of non-adherence, and provides clear explainability when compared to state-of-the-art ML models while remaining competitive on model performance.

Bayesian methods have been applied to model the relationship between adherence and outcomes using dynamic linear models^27,28^, highlighting the time-varying nature of adherence effects. However, these frameworks have not encoded theory- or consumer-informed barrier hierarchies as priors for prospective trajectory prediction from routine primary-care EHRs. Recent GBTM studies in cardiometabolic therapy corroborate the gradual and rapid decline patterns (and variations such as “decline then improve”), yet they do not integrate patient-reported barriers into model specification or produce explanation-ready predictions^8,14,25^. BRIDGE addresses these limitations by embedding consumer- and theory-informed barrier hierarchies directly into a Bayesian predictive framework, enabling prospective prediction of adherence trajectories from routine primary-care EHRs while preserving clinically interpretable links between predicted risk and modifiable barriers. In doing so, the current study contributes a novel barrier-informed, explanation-ready approach that moves beyond descriptive trajectory discovery towards actionable and prospectively deployable adherence prediction.

Finally, counterfactual or recourse analyses are increasingly proposed as a means of linking predictions to actionable interventions. However, health-care-specific guidance emphasises the need for causal constraints, feasibility, and rigorous valuation; generic recourse work also cautions that naïve counterfactuals may not always yield the optimal and equitable outcomes^29^. BRIDGE addresses these concerns by anchoring counterfactual analyses to clinically plausible, barrier-informed levers derived from consumer and theory-based hierarchies, rather than unconstrained feature perturbations. In doing so, the current study contributes an explanation-ready and intervention-oriented framework that links predicted adherence risk to modifiable, contextually feasible targets for care.

### Conceptual advance

BRIDGE integrates behavioural theory with computation by embedding patient-reported barriers as hierarchical priors that inform class-specific coefficients. Although prior trajectory research has convincingly shown heterogeneous, clinically recognisable adherence patterns, most studies have been retrospective and descriptive^8,14^. BRIDGE advances this literature by enabling prospective risk stratification at or near treatment initiation, the point at which intervention is most actionable. To our knowledge, existing Bayesian approaches in adherence research have not incorporated theory- or consumer-informed barrier hierarchies into prior structures, nor have they targeted prediction of future trajectory membership using routinely collected primary care EHR data^28^. Accordingly, we position BRIDGE as a behaviour-informed, interpretable, and deployable prediction framework, rather than a purely data-driven classifier. At the same time, the present study also highlights an important next frontier for behaviour-informed adherence modelling: extending beyond patient and prescribing-history proxies to include clinician-level and consultation-level determinants of adherence. These may include planned time to pathology review, intensity of follow-up, and language used to communicate treatment necessity and expected benefit. Incorporating such features, whether through structured consultation data, prescribing-style indicators, or linked qualitative and survey data, may improve the ability to explain non-initiation and early discontinuation.

The four predicted trajectories, persistent adherence, gradual decline, rapid decline, and early discontinuation, align with archetypes consistently reported across prior trajectory studies^8,10^, our model, BRIDGE seeks to predict at each time point, what are the individualised risk factors that may predict non-adherence and suggest potential clinical interventions that facilitates persistent adherence. Out of the four categories, BRIDGE has elucidated that gradual decline is of particular clinical interest , as it may represent a modifiable intervention window characterised by initial persistence followed by progressive disengagement that might be mitigated if risk is surfaced early. In contrast, rapid decline and early discontinuation suggest narrower windows for intervention, underscoring the importance of early identification and targeted follow-up. BRIDGE preserves temporal heterogeneity, making it more likely to be clinically informative than those that reduce adherence to a single binary indicator or summary metric.

### Actionability via constrained counterfactuals

BRIDGE complements risk estimation with constraint-aware counterfactual analyses that estimate how feasible changes in modifiable factors could shift class membership probabilities towards persistent adherence. Although these analyses are not causal, they provide a structured approach to prioritising intervention levers and align with established behaviour change frameworks, including COM-B and the Behaviour Change Wheel (e.g., education, enablement, environmental restructuring, and treatment review)^30^. In practice, this framework supports the development of actionable decision aids that link individual barrier profiles to feasible, context-specific interventions (e.g., regimen simplification, early follow-up appointments, or proactive management of side effects), while preserving the central role of clinical judgement and patient preferences.

### Methodological transparency and EHR best practice

Consistent with CODE-EHR, we used routinely recorded variables, documented data provenance and curation, and evaluated model performance using multi-class metrics (macro/micro-AUCs, Brier score, log-loss), class-wise reliability, and confusion patterns. We benchmarked performance against strong machine-learning baselines and a parsimonious multinomial regression model to assess added value relative to simpler approaches. A landmark design was implemented to reduces the risk of label leakage. In addition, a baseline-only sensitivity analysis indicated that predictive signal is present at or near treatment initiation.

### Strengths and limitations

This study has several key strengths. First, it integrates patient-reported and theory-informed barriers into a Bayesian hierarchical framework, enabling interpretable and behaviourally grounded prediction of long-term adherence trajectories at or near treatment initiation. Second, by relying solely on routinely collected GP-EMR data, the model is scalable, implementable, and suitable for real-world clinical decision support, while demonstrating robust discrimination, class-level calibration, and quantification of predictive uncertainty. Third, the addition of counterfactual analyses further enhances clinical utility by identifying modifiable levers aligned with patient-centred interventions.

However, several limitations should be considered. The use of EMR-based proxies may not fully capture nuanced psychological, contextual, or prescriber-mediated barriers to adherence. In particular, several potentially important determinants were not directly observed in the GP-EMR data, including the interval planned for pathology review after initiation, the intensity and urgency with which monitoring is framed, and clinicians’ verbal positioning of medicine necessity, benefit, and risk^31^. These factors may influence non-initiation and early discontinuation, but in the present study could only be approximated indirectly through variables such as monitoring recency, visit frequency, prescription timeliness, switching, and biomarker follow-up. Variation in data completeness and measurement variability across practices may also affect model transportability. Although the landmark design minimises the risk of information leakage, some time-varying predictors may still partially reflect emerging adherence behaviour.

## Conclusion

BRIDGE provides a behaviour-informed and interpretable approach for predicting long-term medication-adherence trajectories using routinely collected GP-EMR data. By embedding barrier-informed priors within a transparent Bayesian model, the model offers clinically relevant insights into both risk and underlying drivers of non-adherence. Whith further external validation and prospective evaluation, BRIDGE represents a practical step toward actionable, patient-centred adherence support in primary care.

## Data Availability

The data underlying this article will be shared on reasonable request to the corresponding author and subject to approval by stakeholders.

---

**Supplementary Table S1:** Baseline-only (window 0) model performance.

| Model | AUROC (macro) | G1 | G2 | G3 | G4 | Brier |
| --- | --- | --- | --- | --- | --- | --- |
| *All windows (landmark model)* |  |  |  |  |  |  |
| BRIDGE | 0.808 | 0.710 | 0.939 | 0.762 | 0.830 | 0.531 |
| Random forest | 0.815 | — | — | — | — | 0.548 |
| *Window 0 only (baseline)* |  |  |  |  |  |  |
| BRIDGE | 0.789 | 0.760 | 0.868 | 0.677 | 0.852 | 0.537 |
| Random forest | 0.776 | 0.739 | 0.862 | 0.656 | 0.848 | 0.550 |
| $\Delta$ (BRIDGE) | −0.019 | +0.050 | −0.071 | −0.085 | +0.022 | +0.006 |

To assess temporal circularity, the model was evaluated using only the first six-month observation window per patient, before trajectory-reflective treatment-history patterns have accumulated. Five temporal features (months since initiation and derived interactions) were identically zero and effectively excluded. The modest AUROC decrement (−0.019) confirms that predictive ability is primarily driven by baseline patient characteristics.

**Supplementary Table S2:**
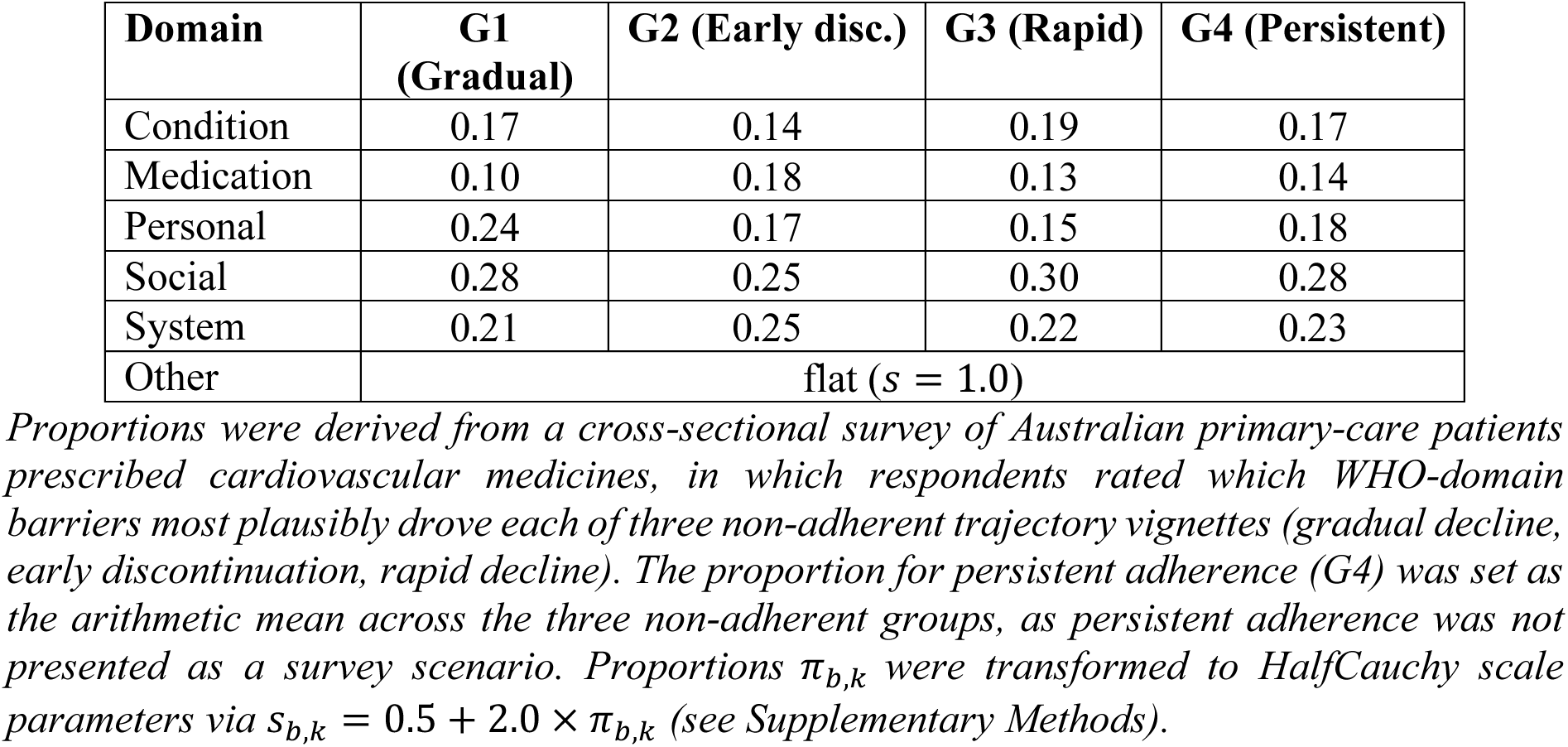
Patient-reported barrier proportions by trajectory group, used to derive barrier-domain hyperprior scales.

**Supplementary Table S3:**
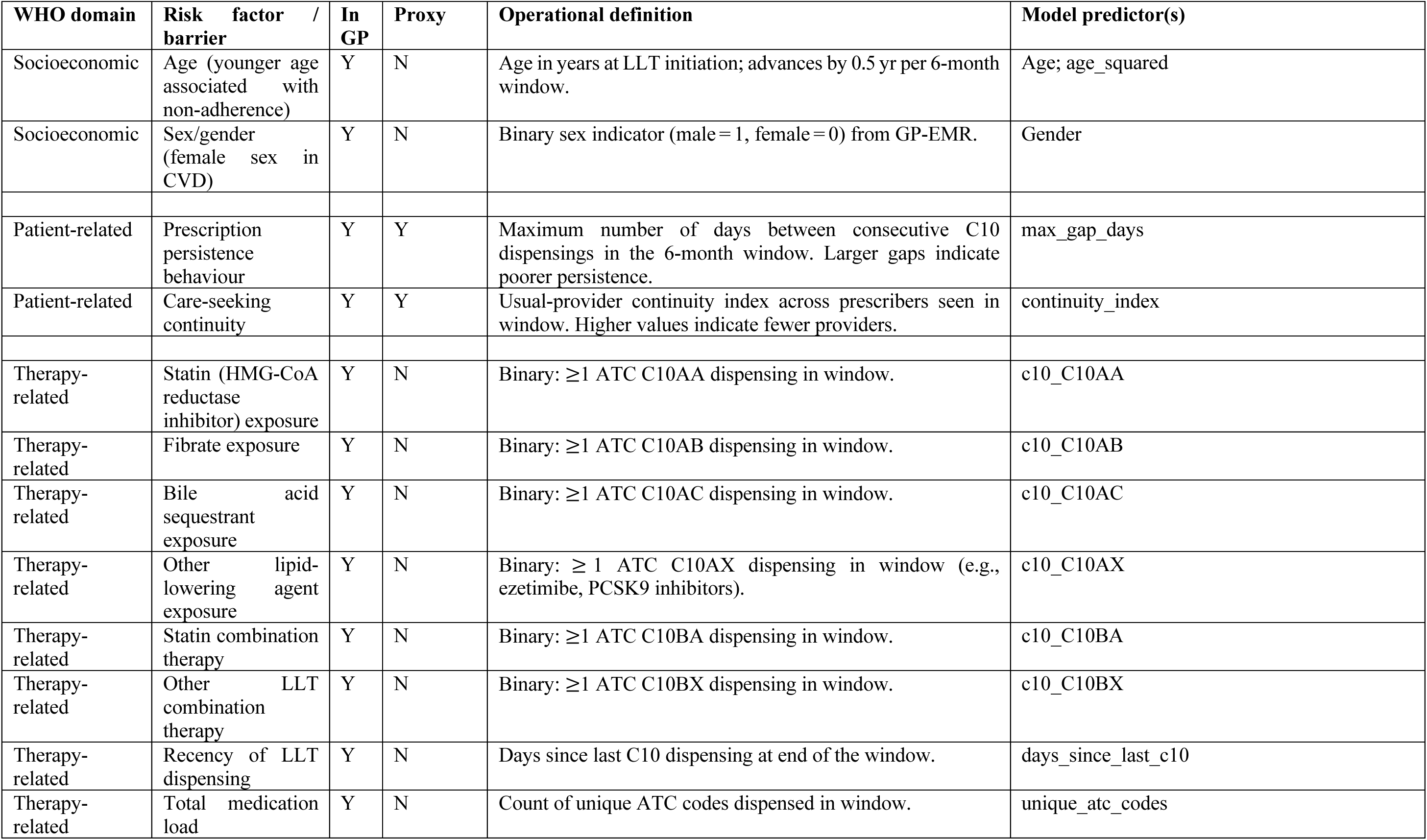
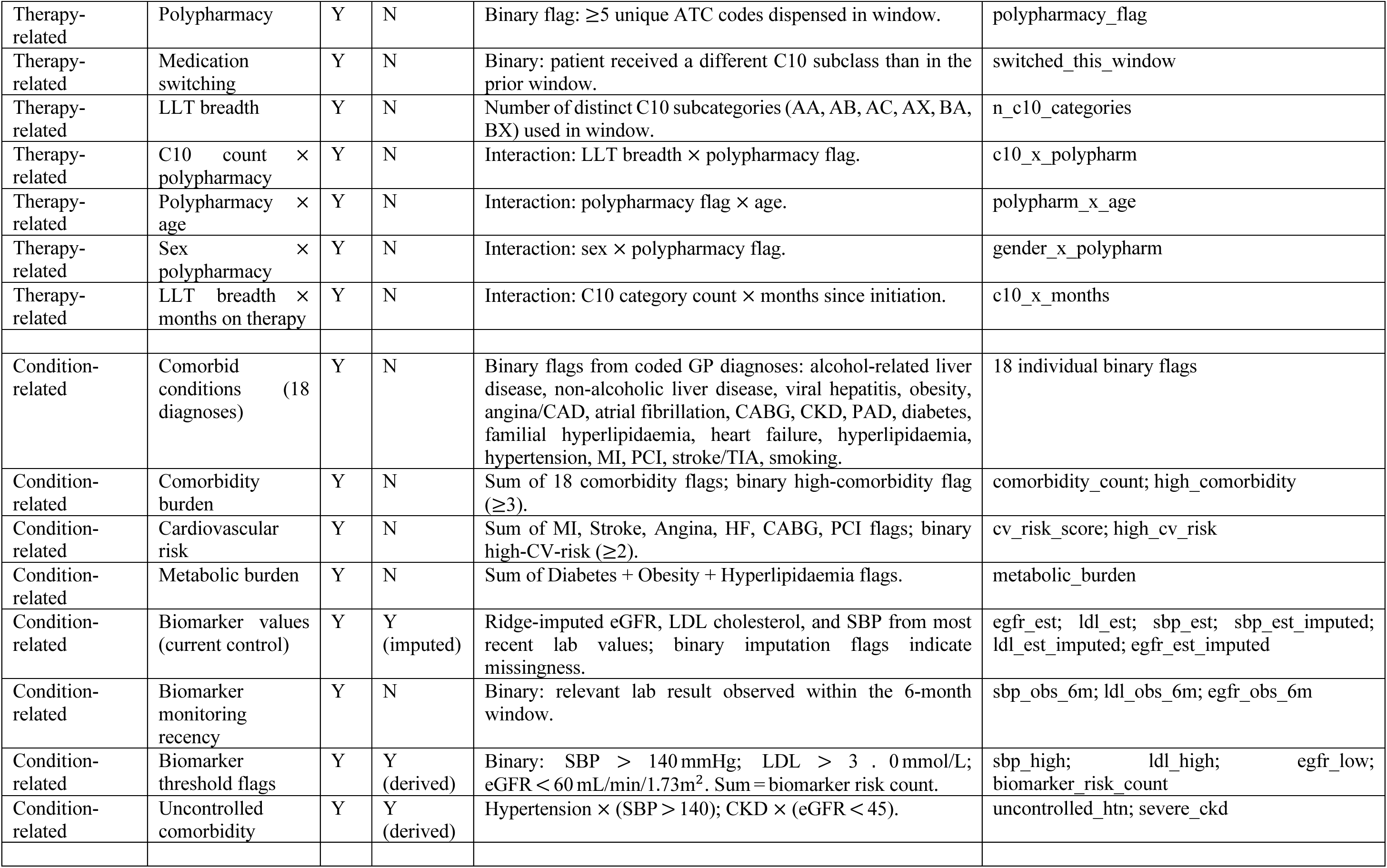
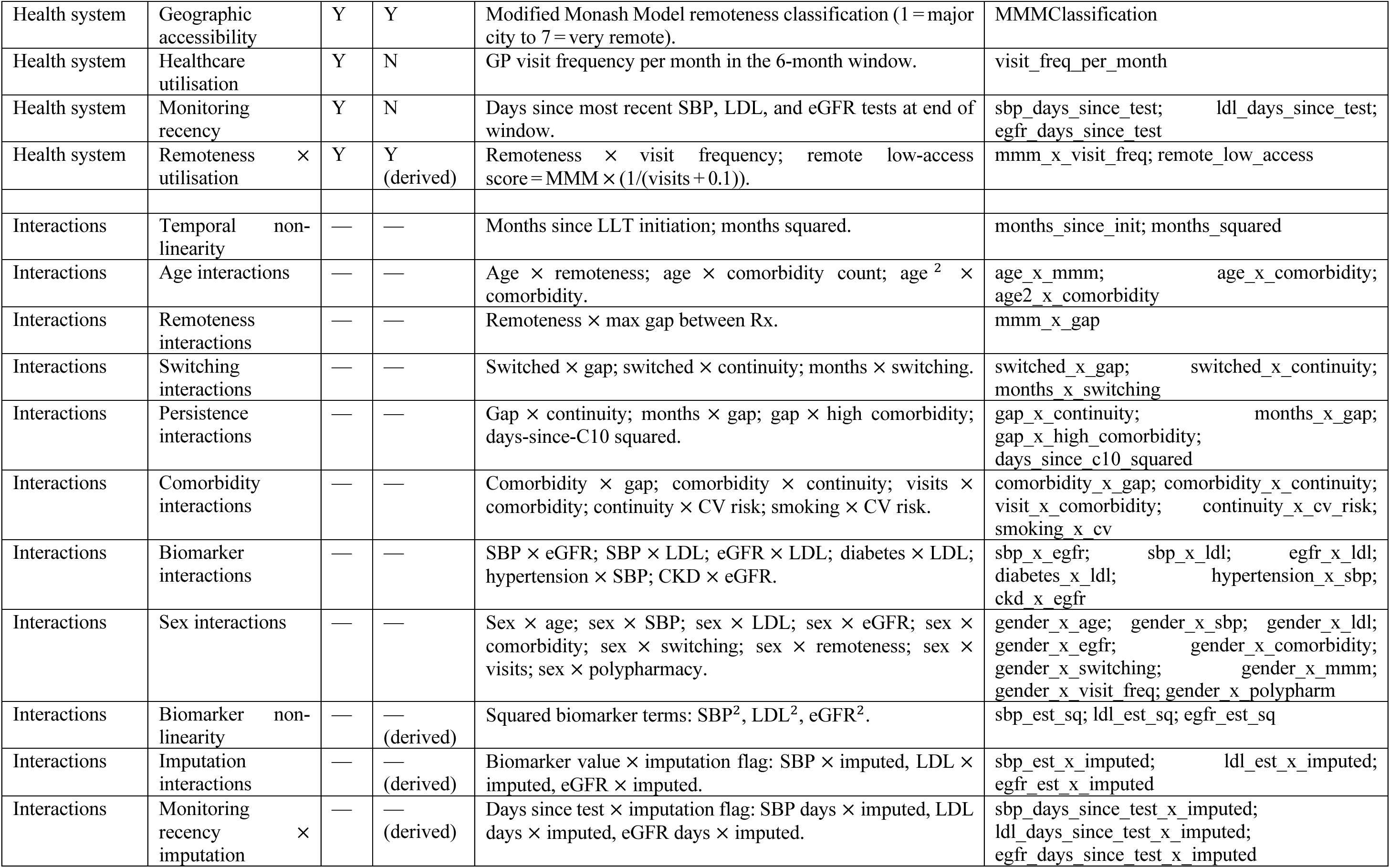
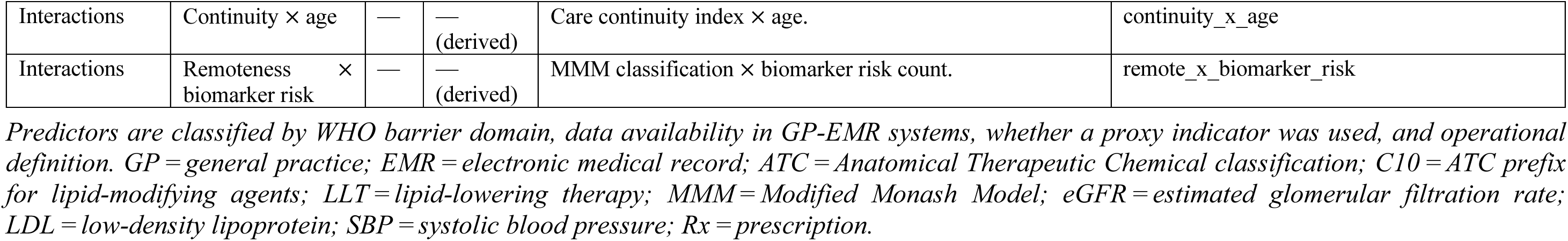
Mapping of model predictors to WHO medication-adherence barrier domains.

**Supplementary Table S4:**
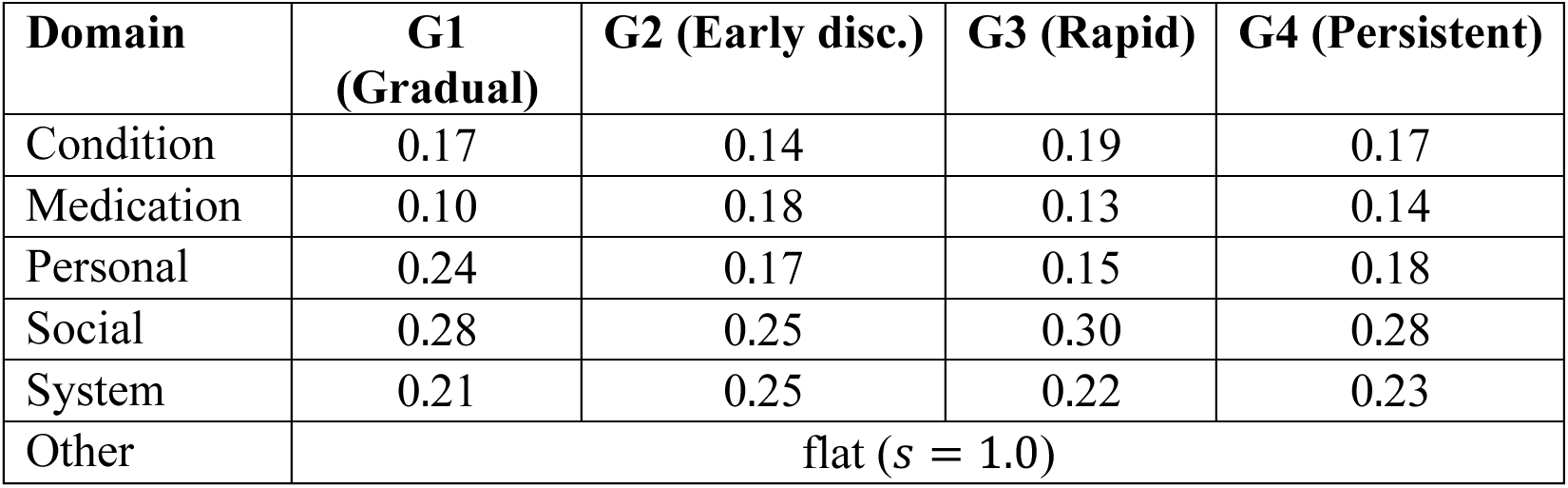
Survey Proportions.

## Supplementary Methods 1: Derivation of Adherence Trajectory Groups Using Group-Based Trajectory Modelling

The four adherence trajectory groups used as outcomes in this study were derived from longitudinal prescription dispensing records using group-based trajectory modelling (GBTM), as previously described. GBTM was applied to rolling 3-month proportion-of-days-covered (PDC) values computed from consecutive prescription fill records over the 36 months following each patient’s index LLT prescription date. PDC per window was calculated as the number of days of LLT supplied divided by the 90-day window length, capped at 1.0.

Models with two to six trajectory groups were evaluated, each specified with a quadratic polynomial time function to accommodate non-monotonic adherence patterns. Group solution selection was based on: (i) Bayesian information criterion (BIC; lower is better), with improvement assessed using the Nagin elbow criterion; (ii) average posterior probability of group membership ≥ 0.70 for all groups, indicating adequate within-group classification certainty; and (iii) minimum group size ≥ 5% of the cohort; and (iv) clinical interpretability assessed by the multidisciplinary study team including pharmacists, general practitioners, and consumer representatives. The four-group solution satisfied all criteria and produced trajectories that were clinically interpretable as distinct non-adherence archetypes.

Trajectory labels were assigned to each patient as their most probable group membership (maximum posterior probability). These static labels served as the outcome variable for all subsequent prediction modelling and group characterisation analyses. The GBTM software used and full model selection statistics (BIC by number of groups, average posterior probabilities, and trajectory shape parameters) are available from the corresponding author on reasonable request.

## Supplemental Methods 2: Survey Methodology for Barrier-Domain Hyperpriors

Patient-reported barrier proportions were elicited through a purpose-designed cross-sectional survey of Australian primary-care patients prescribed cardiovascular medicines (N=708). Respondents were presented with three narrative vignettes representing the three non-adherent trajectory archetypes (gradual decline, early discontinuation, rapid decline) and asked to rate, from a structured list of barrier scenarios drawn from the WHO adherence framework and qualitative synthesis findings, which barriers most plausibly explained each patient’s disengagement from therapy. Barriers were pre-mapped to five WHO adherence domains: patient-related, therapy-related (medication), condition-related, health-system-related, and socioeconomic. The proportion of respondents endorsing each domain for each trajectory vignette was calculated and used to derive the domain-specific HalfCauchy hyperprior scales. The persistent-adherence group (G4) was not presented as a vignette; its proportions were set as the arithmetic mean across the three non-adherent groups, an assumption that persistent adherence is not specifically driven by any single barrier domain. This is a modelling choice rather than an empirical finding, and alternative specifications (e.g., setting G4 proportions to uniform or to the inverse of non-adherent proportions) could produce different regularisation patterns for the reference class. All five barrier domains were directly elicited from survey respondents.

## Supplemental Methods 3: Mathematical Specification of BRIDGE

### Notation

Let 𝑁 denote the number of observation-window rows in the training set, 𝑝 = 108 the number of predictors, and 𝐾 = 4 the number of trajectory classes (G1: gradual decline, G2: early discontinuation, G3: rapid decline, G4: persistent adherence). Let 𝐗 ∈ ℝ*^N^*^×*p*^be the standardised predictor matrix (z-scored to zero mean and unit variance using training-set statistics), 𝐲 ∈ {0,1,2,3}*^N^* the trajectory class labels, and 𝑏(𝑗) ∈ {1, … , 𝐵} a mapping from predictor 𝑗 to one of 𝐵 = 6 WHO-aligned barrier domains (personal, medication, condition, health system, social/demographic, other).

### Survey-derived hyperprior scales

Patient-reported barrier proportions 𝜋*_b,k_* were elicited from a cross-sectional survey of Australian primary-care patients prescribed cardiovascular medicines. For each trajectory class 𝑘 and barrier domain 𝑏, the survey proportion was transformed into a HalfCauchy scale parameter:

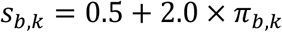

This ensures that barrier domains more strongly endorsed by patients for a given trajectory receive wider prior variance, permitting the corresponding feature weights to take larger values when supported by data. Domains with low survey endorsement receive tighter priors, encoding prior scepticism about their relevance. The “other” domain (interaction terms) received a flat scale of 𝑠 = 1.0. The survey proportions used were:

**Supplementary Table S4a.**
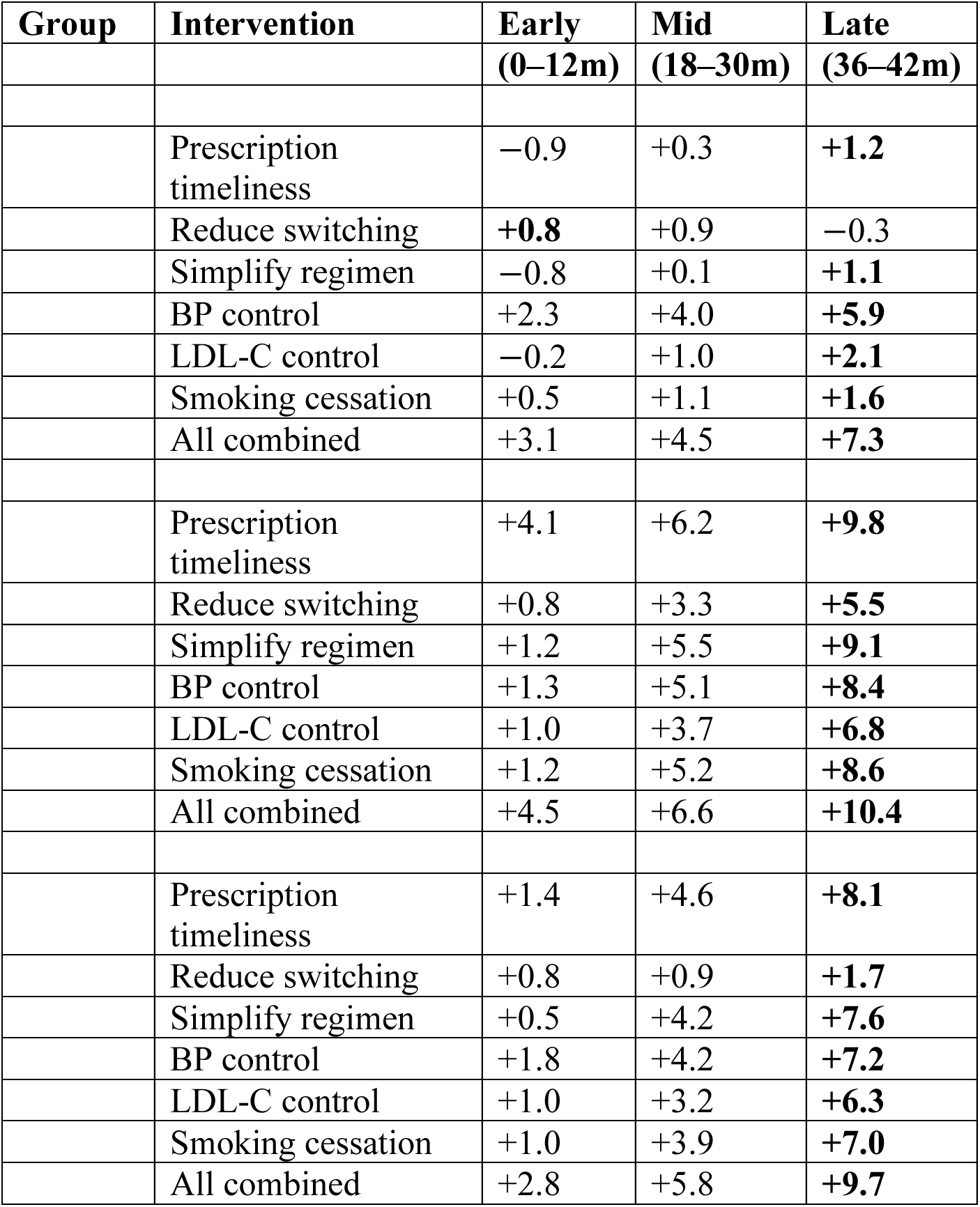
Counterfactual intervention effects by treatment phase. Sample-size-weighted mean change in predicted probability of persistent adherence (𝜟𝑷(G4), percentage points) for each intervention lever, aggregated across early (0–12 months), mid (18–30 months), and late (36–42 months) observation windows. Values derived from nearest-neighbour counterfactual matching (𝒌 = 𝟏𝟎). Bold values indicate the phase with the largest effect for each group–lever combination. Full per-window results are in the extended version of this table (available as supplementary data).

### Generative model

The full generative model is specified as follows:

**Level 1 — Barrier-domain variance parameters.** For each barrier domain 𝑏 ∈ {1, … ,6} and class 𝑘 ∈ {1, … ,4}:

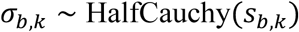

These 6 × 4 = 24 variance parameters control domain- and class-specific regularisation strength.

**Level 2 — Feature weights.** For each predictor 𝑗 ∈ {1, … ,108} and class 𝑘 ∈ {1, … ,4}:

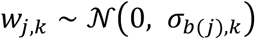

where 𝑏(𝑗) maps predictor 𝑗 to its assigned barrier domain. This induces partial pooling: predictors within the same domain share a common variance parameter, but the variance is allowed to differ by trajectory class. Domains with tight posterior 𝜎 values indicate that the data did not support large effects for that domain–class combination.

### Level 2 — Bias terms

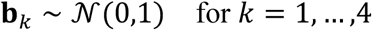

**Likelihood — Standard categorical (no class weighting).** For each observation 𝑖:

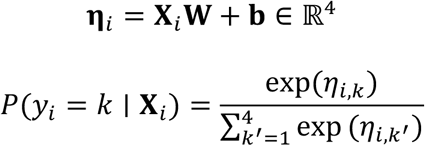

The production model uses a standard (unweighted) categorical log-likelihood:

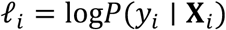

A class-weighted variant was evaluated (inverse-frequency weighting 𝜔*_k_* = *N*/(4*N_k_*)) but yielded lower AUROC and worse calibration (Supplementary Table S5), so the unweighted likelihood was retained. The barrier-informed hierarchical priors provide implicit regularisation across trajectory classes with varying prevalences (10–41%).

### Inference

Parameters were estimated via stochastic variational inference (SVI) using a mean-field (diagonal normal) variational family:

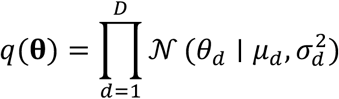

where 𝛉 = {𝛔_barrier_, 𝐖, 𝐛} collects all latent variables and 𝐷 is the total number of parameters. The variational parameters {𝜇_1_, 𝜎_1_} were optimised by minimising the evidence lower bound (ELBO) loss using the Adam optimiser (learning rate = 0.02) over 100 000 steps. The fitted variational posterior 𝑞(𝛉) can be sampled to obtain draws for downstream uncertainty quantification.

### Prediction

At test time, point predictions use the variational posterior mean weights ̂**W** = E_q_[**W**] and bias ̂**b** = E_q_[**b**]:

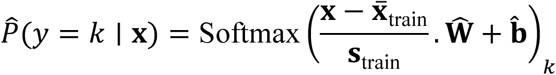

where ̄X_train_ and S_train_ are the training-set feature means and standard deviations used for z-score standardisation. For uncertainty quantification, predictions are computed over 2 000 draws from the fitted variational posterior to obtain credible intervals on predicted probabilities and derived quantities (e.g., barrier-domain variance parameters, feature-weight distributions; see Supplementary Figure S4).

### Implementation

The model was implemented using NumPyro with JAX for GPU-accelerated computation. The variational guide was AutoDiagonalNormal. Feature standardisation, interaction-term construction, and post-hoc analyses (partial-dependence grids, counterfactual simulations) were performed in NumPy/Pandas on CPU.

### Panel C: Medication Simplification Scenario Definitions

Six medication simplification scenarios were evaluated in the counterfactual analysis:

1. **Reduce total drug types**: cap unique ATC codes at the comorbidity-aware minimum.
2. **Remove polypharmacy**: reduce total unique ATC codes to below the polypharmacy threshold (<5 concurrent drugs), respecting the comorbidity floor, and recalculate the polypharmacy flag. Only patients whose comorbidity floor was below 5 were eligible; patients whose essential medication needs already required ≥ 5 drugs were left unchanged.
3. **Consolidate to single LLT class**: reduce to one lipid-lowering drug category.
4. **Lipid-lowering polypill**: combine multiple C10 subcategories into a single fixed-dose combination, reducing total drug count and recalculating the polypharmacy flag.
5. **Statin-only therapy**: single LLT class, comorbidity-floored ATC codes, statin (C10AA) indicator set to 1, all other C10 subcategory indicators set to 0.
6. **Deprescribe two drugs**: reduce unique ATC codes by two (floored at comorbidity minimum) and remove the polypharmacy flag.

All scenarios respected a per-patient comorbidity-aware minimum: one drug for LLT plus one additional drug for each of diabetes, hypertension, heart failure, chronic kidney disease, stroke, and peripheral arterial disease, reflecting the minimum chronic pharmacotherapy required by each condition. Patients already at or below their comorbidity floor were unaffected. All drug-count operations produced whole-number values (via floor rounding). Each scenario was evaluated at both the population level and within the polypharmacy subgroup (≥5 concurrent medications). Temporal interaction terms were frozen as in Panel B.

## Supplementary Figures

**Supplementary Figure S1.**
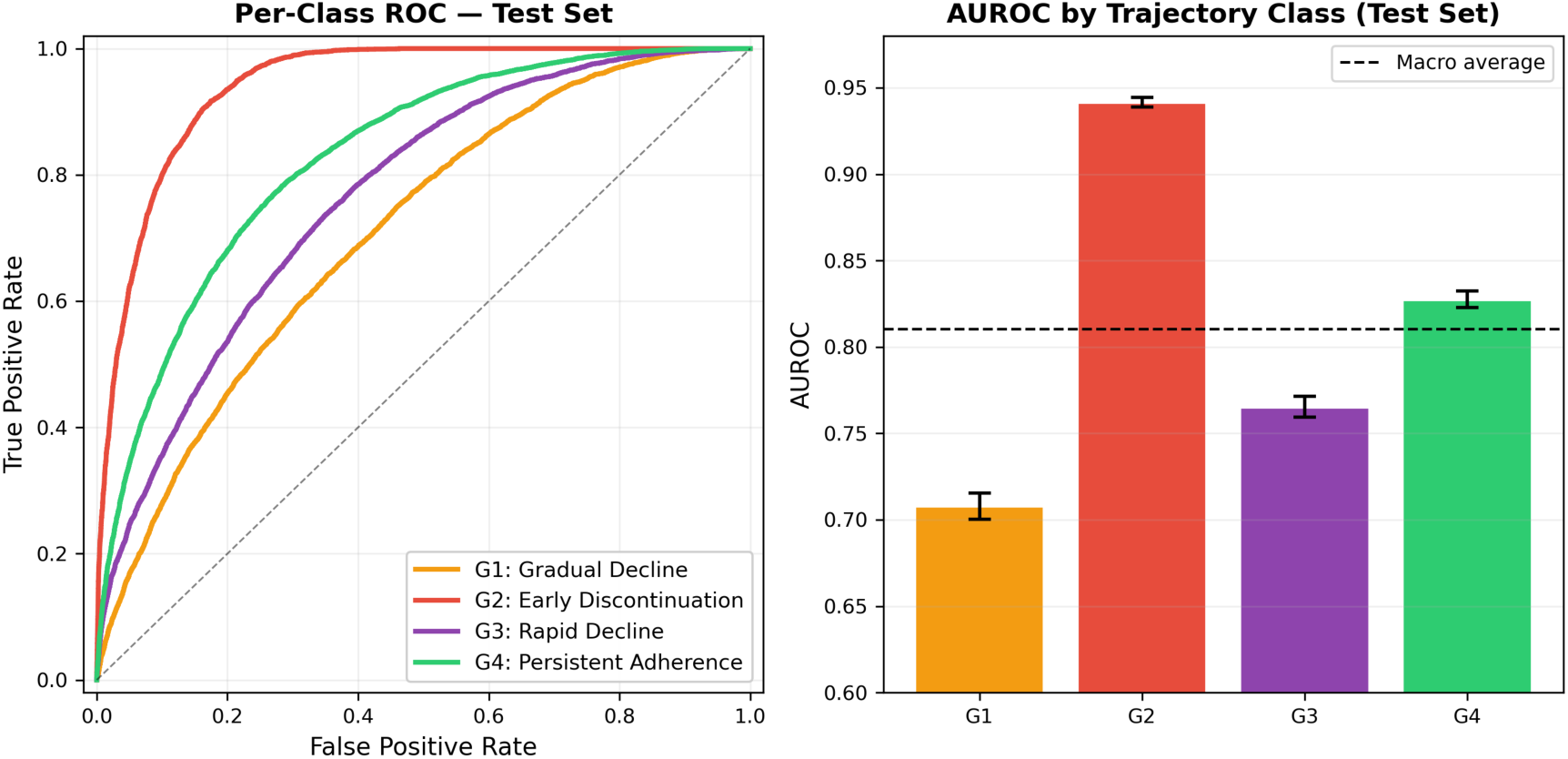
Held-out test-set per-class ROC curves. One-vs-rest AUROC is shown for each trajectory class on the fixed test cohort, with 95% bootstrap confidence intervals.

**Supplementary Figure S2.**
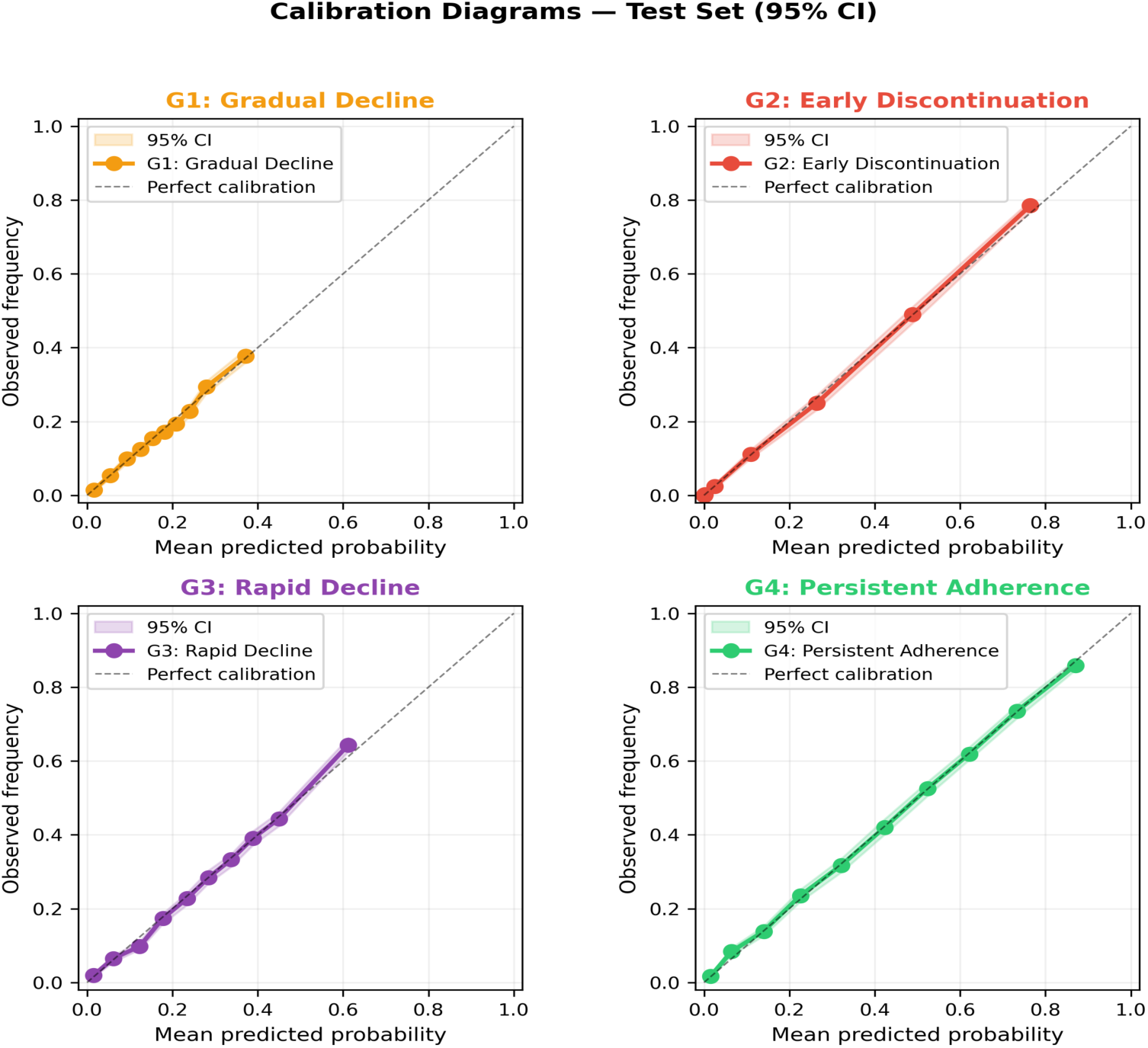
Calibration (reliability) diagrams by trajectory class: Each panel shows the relationship between mean predicted probability and observed frequency for one trajectory class, using quantile-based binning (10 bins). Points close to the diagonal indicate good calibration. Per-class Brier scores: G1 = 0.13, G2 = 0.07, G3 = 0.16, G4 = 0.16. Calibration is good across all classes, with the shaded area indicating deviation from perfect calibration.

**Supplementary Figure S3.**
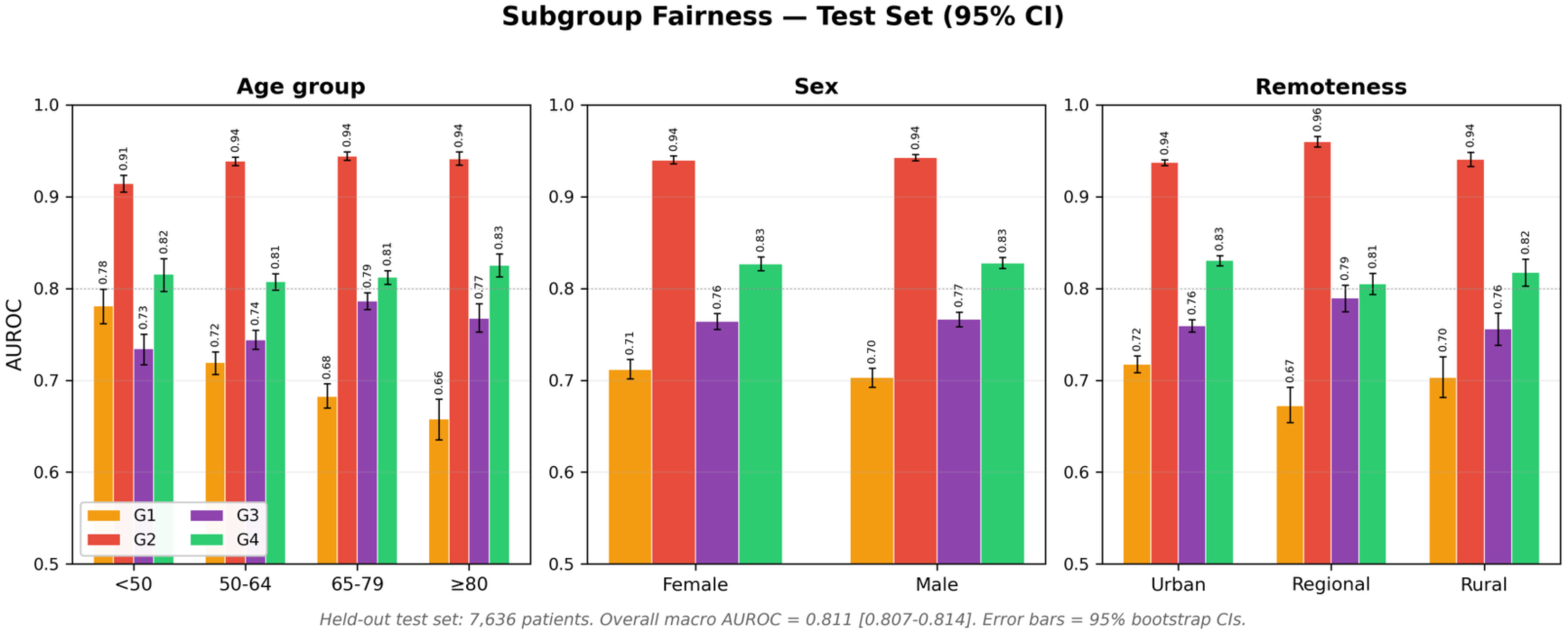
Held-out test-set subgroup fairness. Macro and per-class AUROC are reported across age, sex, and remoteness strata in the fixed test cohort.

**Supplementary Figure S4.**
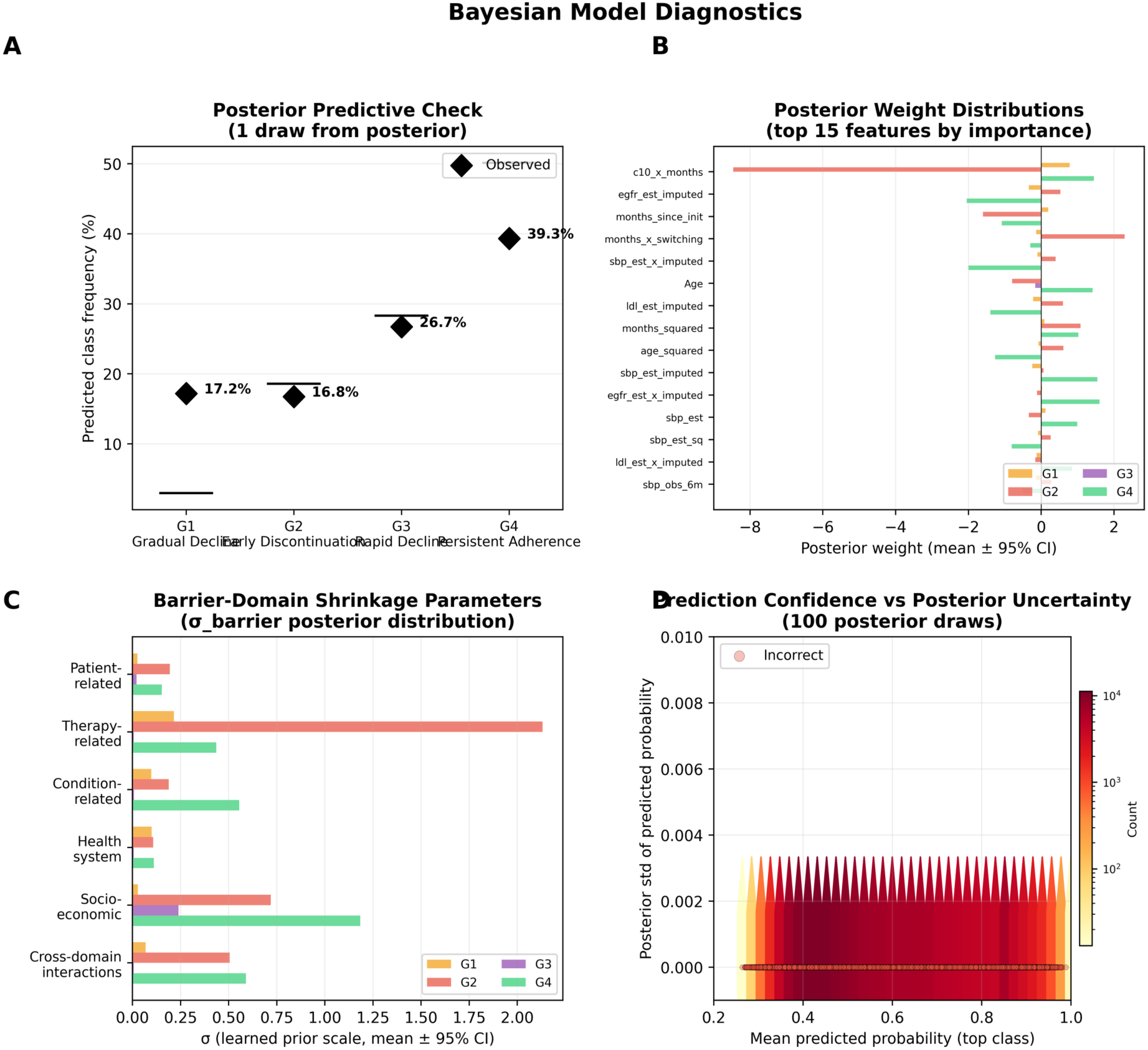
Bayesian model diagnostics (500 posterior draws, test set 𝒏 = 28 107 observations). Panel A: Posterior predictive check comparing observed test-set class frequencies (black diamonds) with distributions predicted by 200 posterior draws (box plots). All four classes are well calibrated: G1 observed 17% vs predicted median 17% [17–18%]; G2 17% vs 17% [16–17%]; G3 27% vs 27% [26–28%]; G4 39% vs 39% [39–40%]. The posterior predictive distribution closely recovers the observed class prevalences, confirming adequate model specification. Panel B: Posterior barrier-domain variance parameters ( 𝝈_barrier_ ) with 95% credible intervals. Larger 𝝈 permits greater deviation from the prior mean, indicating domains where the data provide strong signal. G2 (early discontinuation) predictions are dominated by therapy-related barriers (𝝈 = 2.1), while G3 (rapid decline) features are tightly regularised across most domains ( 𝝈 < 0 . 01 for therapy and condition), with socioeconomic factors (𝝈 = 0.29) permitted greater flexibility. G4 (persistent) shows the largest socioeconomic variance (𝝈 = 0.93), reflecting the strong age and sex effects in sustained adherence. Panel C: Posterior weight distributions for the 15 most influential features, showing medians with 95% credible intervals per class. Features with narrow CIs excluding zero (e.g., c10_x_months for G2, Age for G4) represent high-confidence predictors; wide CIs reflect greater posterior uncertainty. Panel D: Model summary statistics, including MAP log-likelihood (−26 273), posterior mean log-likelihood (−26 312, SD 22), and MAP accuracy (59%). Parameters were estimated via stochastic variational inference with a diagonal normal guide over 100 000 steps.

**Supplementary Figure S5.**
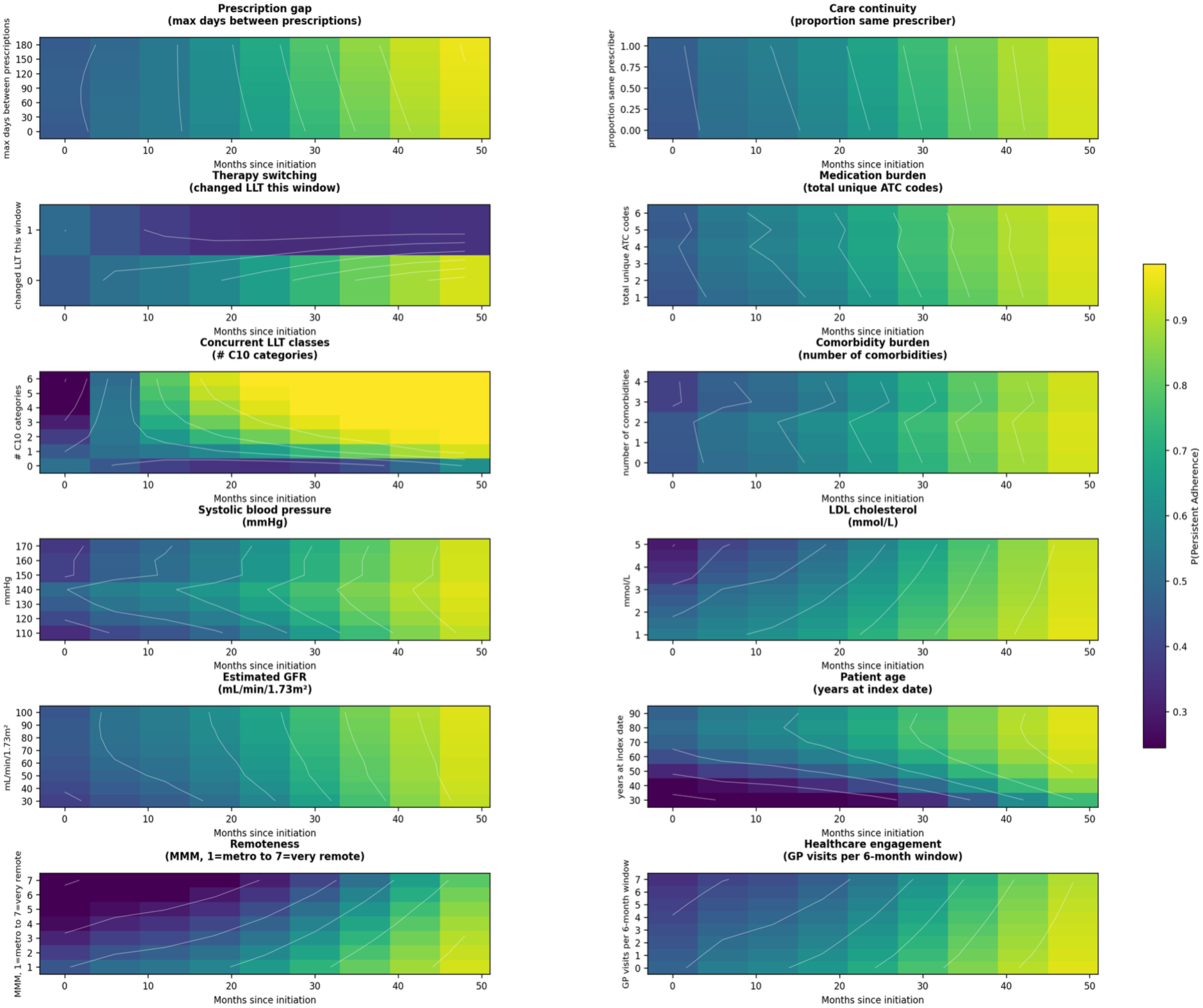
Extended partial-dependence analysis. Partial-dependence grids for 12 clinical dimensions showing P(Persistent Adherence) as a function of each factor (y-axis) over months since LLT initiation (x-axis), with all other predictors held at test-set median. Panels include prescribing factors (prescription gap, care continuity, therapy switching, medication burden, concurrent LLT classes), biomarkers (systolic blood pressure, LDL cholesterol, estimated GFR), demographics (age, remoteness), and healthcare engagement (GP visits).

**Supplementary Figure S6.**
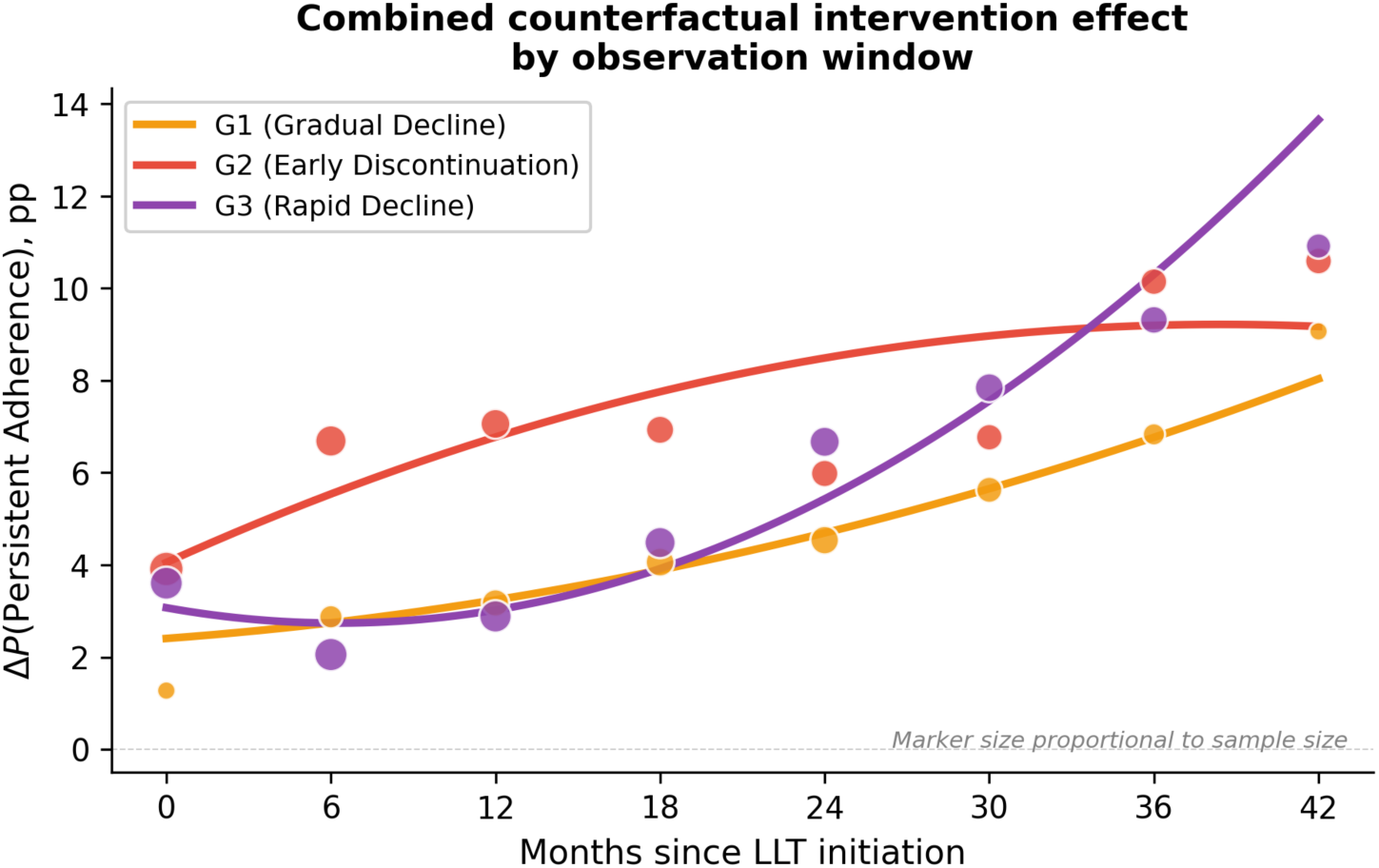
Combined counterfactual intervention effect by observation window. Sample-size-weighted quadratic trend (solid line) of the combined nearest-neighbour counterfactual 𝜟𝑷(G4) across eight six-month observation windows for each non-adherent trajectory group. Marker size is proportional to sample size at each window. Intervention effects grow monotonically with time on therapy for all three groups, consistent with progressive behavioural divergence from persistent adherers creating larger scope for modifiable-feature substitution at later windows.

**Supplementary Figure S7.**
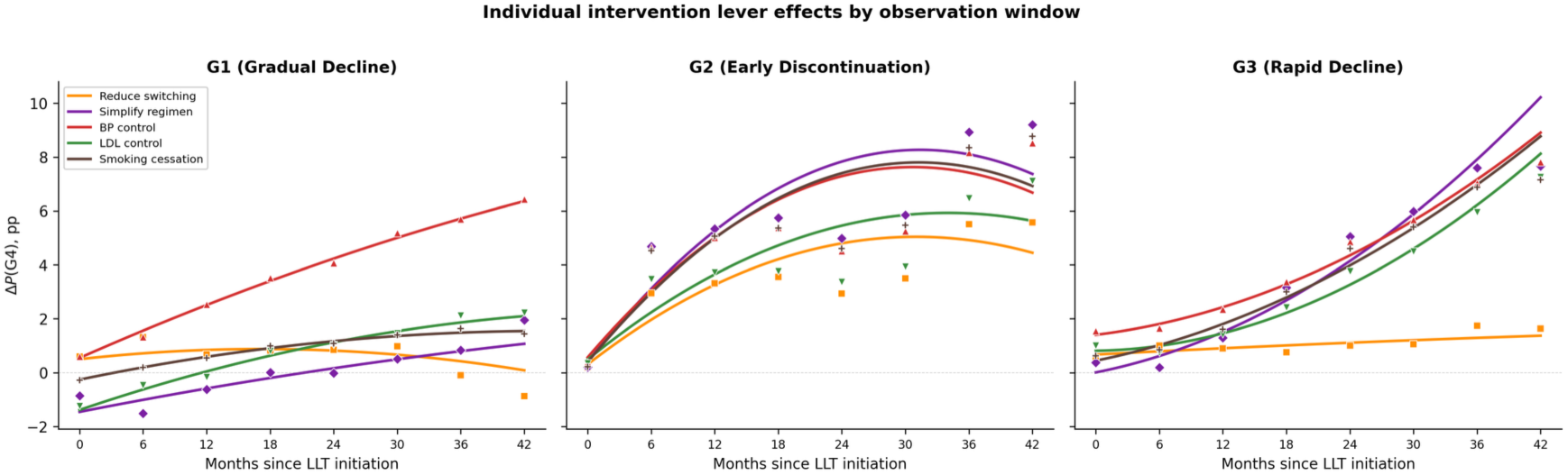
Individual intervention lever effects by observation window. Sample-size-weighted quadratic trends for each of six intervention levers across eight observation windows, shown separately for G1 (gradual decline), G2 (early discontinuation), and G3 (rapid decline). For G1, blood pressure control is the dominant lever from 12 months onward. For G2, prescription timeliness dominates across all windows. For G3, all levers converge at similar magnitudes by late windows.

